# Cohort monitoring of 29 Adverse Events of Special Interest prior to and after COVID-19 vaccination in four large European electronic healthcare data sources

**DOI:** 10.1101/2022.08.17.22278894

**Authors:** Miriam Sturkenboom, Davide Messina, Olga Paoletti, Airam de Burgos-Gonzalez, Patricia García-Poza, Consuelo Huerta, Ana Llorente- García, Mar Martin-Perez, Maria Martinez, Ivonne Martin, Jetty Overbeek, Marc Padros-Goossens, Patrick Souverein, Karin Swart, Olaf Klungel, Rosa Gini

## Abstract

**Setting:** Primary and/or secondary health care data from four European countries: Italy, the Netherlands, the United Kingdom, Spain

**Participants:** Individuals with complete data for the year preceding enrollment or those born at the start of observation time. The cohort comprised 25,720,158 subjects.

**Interventions:** First and second dose of Pfizer, AstraZeneca, Moderna, or Janssen COVID-19 vaccine.

**Main outcome measures:** 29 adverse events of special interest (AESI): acute aseptic arthritis, acute coronary artery disease, acute disseminated encephalomyelitis (ADEM), acute kidney injury, acute liver injury, acute respiratory distress syndrome, anaphylaxis, anosmia or ageusia, arrhythmia, Bells’ palsy, chilblain-like lesions death, erythema multiforme, Guillain Barré Syndrome (GBS), generalized convulsion, haemorrhagic stroke, heart failure, ischemic stroke, meningoencephalitis, microangiopathy, multisystem inflammatory syndrome, myo/pericarditis, myocarditis, narcolepsy, single organ cutaneous vasculitis (SOCV), stress cardiomyopathy, thrombocytopenia, thrombotic thrombocytopenia syndrome (TTS) venous thromboembolism (VTE)

**Results:** 12,117,458 individuals received at least a first dose of COVID-19 vaccine: 54% with Comirnaty (Pfizer), 6% Spikevax (Moderna), 38% Vaxzevria (AstraZeneca) and 2% Janssen Covid-19 vaccine. AESI were very rare <10/100,000 PY in 2020, only thrombotic and cardiac events were uncommon. After adjustment for factors associated with severe COVID, 10 statistically significant associations of pooled incidence rate ratios remained based on dose 1 and 2 combined. These comprised anaphylaxis after AstraZeneca vaccine, TTS after both AstraZeneca and Janssen vaccine, erythema multiforme after Moderna, GBS after Janssen vaccine, SOCV after Janssen vaccine, thrombocytopenia after Janssen and Moderna vaccine and VTE after Moderna and Pfizer vaccines. The pooled rate ratio was more than two-fold increased only for TTS, SOCV and thrombocytopenia.

**Conclusion:** We showed associations with several AESI, which remained after adjustment for factors that determined vaccine roll out. Hypotheses testing studies are required to establish causality.

## Introduction

The global and rapid spread of COVID-19 caused by the SARS-CoV2 triggered the need for developing vaccines to control for this pandemic[1]. COVID-19 vaccine development has been triggered on a global level following the release of the genetic sequence of SARS-CoV2 on 11 January 2020[2]. The landscape for COVID-19 vaccines is characterized by a wide range of technology platforms including nucleic acid (DNA and RNA), virus-like particle, peptide, viral vector (replicating and non-replicating), recombinant protein, live attenuated virus and inactivated virus approaches. The world has seen an unprecedented speed in which the vaccines were rolled out, with first emergency authorizations of COVID-19 vaccines, within a year after the identification of the genetic sequence. In Europe, 5 different vaccines have been conditionally authorized including Comirnaty (Pfizer/BioNtech, December 2020), Vaxzevria (AstraZeneca, January 2021), Spikevax (Moderna, February 2021) Covid-19 vaccine Janssen (March 2021) and Nuvaxovid (Novavax, December 2021)[3].

Due to the rapid development of new COVID-19 vaccines, many questions were raised about the benefits and risks for the vaccines at individual and population levels[4]. Several emerging safety signals of rare serious events have been detected after COVID-19 vaccine launch on the basis of case reports or case series, such as anaphylaxis[5-7], thrombotic events following vaccines with adenovirus platforms[8-11], the rare occurrence of vaccine induced thrombotic thrombocytopenia, thrombocytopenia itself, capillary leak syndrome[12, 13], myocarditis[14-16] and Guillain-Barre Syndrome[17]

The EU safety monitoring plan for COVID-19 vaccines requires the European Medicines Agency (EMA) to monitor suspected side effects reported by individuals and healthcare professionals in the EU [18]. An EU database called EudraVigilance holds these reports. EMA’s Pharmacovigilance Risk Assessment Committee (PRAC) and the national competent authorities (national regulatory agencies) in the EU continuously monitor EudraVigilance to identify any new safety issues that require investigation. These are known as safety signals. When assessing a safety signal, the PRAC looks for any unusual or unexpected patterns, such as a medical event occurring in vaccinated people at a higher rate than in the general population, a description of the regulatory monitoring of covid-19 vaccines process is available online[19].

The Early Covid Vaccine Monitoring (ECVM) study was funded by the EMA[20] and complements the above-mentioned passive surveillance system. The ECVM project comprised of two distinct parts: a prospective cohort event monitoring based on patient reported data in 6 countries, and a cohort monitoring of exposure, adverse events of special interest (including coagulation disorders) in vaccinated persons (by brand) using 4 electronic healthcare data sources covering populations in four countries (the Netherlands, Italy, Spain and the United Kingdom) prior to and after COVID-19 vaccination. In this paper we report on the cohort monitoring of AESI using 4 electronic health care data according to the protocol that is available publicly in the EU PAS register[21]. Objectives were to monitor and estimate the incidence rates of adverse events of special interest (AESI) in vaccinated and non-vaccinated persons by data source over the period January 1st 2020-October 31^st^ 2021 by brand and dose of vaccine and to compare the incidence rates of AESIs in the window 28 days after vaccination with dose 1 or dose 2 with the incidence rates of AESIs of non-vaccinated people in 2020. This study was not designed for causal inference. The protocol is publicly available (EUPAS 40404).

## Methods

### Design & population

We performed a cross-national multi-database retrospective dynamic cohort study, in adult and paediatric individuals from January 2020, 1^st^ to October 2021, 31^st^, or until the date of last data available in the data source.

Individuals were required to have at least 365 days of data availability before cohort entry, except for those who entered the cohort at birth. The end of follow-up was the earliest dates of event occurrence (specific per AESI), last data collection or death.

In the cohort study person-time after start of study is divided in two main periods, non-vaccinated time, which is from 1/1/2020 until the moment of first COVID-19 vaccination and vaccinated person-time, which starts at the first of any of the COVID-19 vaccine, and lasts for a maximum of 28 days after dose 1 and 28 days after dose 2. To avoid confounding by vaccine roll-out strategies non-vaccinated time was limited to 2020.

### Setting

The study includes hospital and outpatient data from electronic healthcare data sources in 4 different European countries, which were chosen since they have short data lag times and capture COVID-19 vaccines. Details have been described before in the ACCESS study [22], also all data sources are listed on the ENCePP public catalogue of data sources. In summary 1) the PHARMO Database Network from the Netherlands, which for this analysis used information from general practice medical records in the Netherlands. The records include information on diagnoses and symptoms, laboratory test results, referrals to specialists and healthcare product/drug prescriptions[23]. 2) the BIFAP (Base de Datos para la Investigación Farmacoepidemiológica en Atención Primaria (BIFAP) database[24], comprises medical records of primary care. Additionally, information on hospital discharge diagnoses is linked to patients included in BIFAP for a subset of periods and regions participating in the database. BIFAP has been linked to a COVID registry for COVID monitoring. The data from all these settings in BIFAP has been used for the current analysis but was split into subpopulations of GP data and GP plus hospitalizations; 3) the Italian database maintained by the Tuscan regional office (ARS) comprises all data banks that are collected by the Tuscany Region to account for the healthcare delivered to its around 3.6 million inhabitants. It routinely collects primary care and secondary care prescriptions of drugs for outpatient use and can link these databanks at the individual level with hospital admissions, and admissions to emergency care, records of exemptions from co-payment, diagnostic tests and procedures, causes of death, mental health services registry, birth registry, spontaneous abortion registry, induced terminations registry, COVID registry[25]; 4) the Clinical Practice Research Datalink (CPRD) from the United Kingdom collates the computerized medical records of GPs. The data recorded include demographic information, prescription details, clinical events, preventive care, specialist referrals, hospital admissions, and major outcomes, including death. For this analysis, we used CPRD Aurum (June2021)[26]. Use of CPRD data for this project was approved by CPRD’s Research Data Governance (RDG) Process (protocol no. 21_000429). The use of PHARMO data was approved by the Institutional Review Board of ‘Stichting Informatie voorziening voor Zorg en Onderzoek’ (STIZON, ID CC2021-21). Use of BIFAP data for this project was approved by the Scientific Committee of BIFAP (protocol reference: 01/2021) and an Ethics Committee (Comité de Ética de La Investigación con Medicamentos del Hospital Universitario de la Princesa: aprobación 11-03-21, acta CEIm 05/21). A summary of the vocabularies used for the diagnosis codes across these data sources are provided in Table 1.

**Table 1:** Characteristics of participating data sources and attrition.

|  | **Italy_ARS** | **ES_BIFAP** | **ES_BIFAP** | **NL_PHARMO** | **UK_CPRD-Aurum** |
| --- | --- | --- | --- | --- | --- |
| Type of data sources | Record linkage | Medical records primary care | Medical records primary care linked to hospitalization discharge | Medical records primary care | Medical records primary care |
| Data end date (may be earlier than actual end to calendar date of data extraction to make sure diagnoses are covered) | 30jun2021 | 31aug2021 | 31aug2021 | 01aug2021 | 09may2021 |
| Diagnosis vocabularies | ICD9CM | ICPC, SNOMED | SNOMED, ICD10CM | ICPC | SNOMED/MedCodeID, RCD2 |
| **Attrition** |  |  |  |  |  |
| Persons in the instance of the data source | 3,780,912 | 7,780,289 | 7,780,289 | 2,343,154 | 17,235,829 |
| Sex or birth date missing or absurd, no dates of entry or exit | 0 | 10 | 10 | 7 | 2466 |
| Death before study start | 49071 | 550903 | 550903 | 0 | 0 |
| Exit from the data source before study start | 201310 | 1283926 | 4143292 | 1446 | 1862872 |
| Persons in the data source at study start | 3530531 | 5945450 | 3086084 | 2341701 | 15370491 |
| Less than 365 days history at 1//1/2020 | 40908 | 128895 | 51158 | 29247 | 1268965 |
| Study population | **3,489,623** | **5,816,555** | **3,034,926** | **2,312,454** | **14,101,526** |

### Adverse Events of Special Interest (AESI)

The list of AESI was based on the initial list of AESI that was defined by the Coalition for Epidemic Preparedness Innovations (CEPI) funded SPEAC project conducted by the Brighton Collaboration. The list has been defined based on events that are related or potentially related to marketed vaccines, events related to vaccine platforms or adjuvants, and events that may be associated with COVID-19[27]. This preliminary list was extended for the EMA-funded vACcine Covid-19 monitoring readinESS (ACCESS) project [28] and was agreed with the EMA advisory group monitoring committee[22]. Bell’s palsy was added to the initial ACCESS AESI list for this ECVM study.

The list of 29 AESIs comprised in alphabetical order: acute aseptic arthritis[29], acute coronary artery disease[30], acute disseminated encephalomyelitis (ADEM)[31], acute kidney injury[32], acute liver injury[33], acute respiratory distress syndrome[34], anaphylaxis[35], anosmia or ageusia[36], arrhythmia[37], Bells’ palsy, chilblain-like lesions[38], death (defined as date of death in persons table), erythema multiforme[39], Guillain Barré Syndrome (GBS)[40], generalized convulsion[41], haemorrhagic stroke[42], heart failure[43], ischemic stroke[42], meningoencephalitis[44], microangiopathy[45], multisystem inflammatory syndrome[46], myo/pericarditis[47], myocarditis[47], narcolepsy[48], single organ cutaneous vasculitis (SOCV)[49], stress cardiomyopathy[50], thrombocytopenia[51], thrombotic thrombocytopenia syndrome (defined as arterial or venous thrombotic events & thrombocytopenia within 10 days)[42], venous thromboembolism (VTE)[42]. Sudden death and diabetes mellitus 1, which were in the protocol could not be studied adequately without further validation of algorithms and are not included in this publication. For acute aseptic arthritis no specific codes could be identified in ICD vocabularies.

To create the outcome variables, semantic harmonization was required in order to manage the heterogeneity in diagnosis coding terminologies across DAPs. The first step was conducted using the Codemapper[52], which allows for semi-automated mapping on the basis of the Unified Medical Language System (UMLS) across the different diagnosis terminologies that are used by the DAPs: the International Classification of Diseases, Ninth Revision, Clinical Modification (ICD-9-CM), International Classification for Primary Care (ICPC) and ICD Tenth Revision (ICD-10-CM), Readv2, SNOMED clinical CT US Edition and Spanish Edition (SCTSPA). Codemapper allows for tagging of concepts (e.g. narrow (specific) or broad (sensitive) concepts) and produces a code list. The code list and tagging was reviewed by all DAPs. Only specific (narrow) codes were included in the common analysis script to create the study variables, to reduce misclassification.

The date of an event was the first occurrence of a record of a diagnostic code for such an event during follow-up, except for anaphylaxis and general convulsions which could be repeated. None of the events was validated because of time and limited budget. When events could not be extracted reliably DAPs had the right to ask to not present these events, this happened for DM1, which was mixed with diabetes mellitus type 2 in several sites and with sudden death, which could not be identified without proper validation, for BIFAP transverse myelitis is not shown.

### Exposure to COVID-19 vaccinations

Exposure was based on any type of record that contained a dispensing or administration of any of the authorized COVID-19 vaccines. Vaccine manufacturer and date of vaccination were obtained from all data sources. As Comirnaty, Spikevax and Vaxzevria are all licensed as a two-dose primary vaccination series, multiple vaccinations per person were identified. Exposure to these vaccines was classified by brand, dose and week of administration and counted for exposure monitoring. When dose was not available this was inferred by ordering the vaccinations by date for a person, and the second dose was required to be at least 14 days distant from the first and the third at least 90 days from the second. All entries of a covid vaccine within 14 days after the first dose for a person were deleted. This cleaning step was necessary as some data sources receive notification of covid-vaccination through multiple routes (e. g. PHARMO).

### Covariates

Covariates comprised factors that are associated with roll-out of the vaccination campaign: age, gender, prior COVID-19 diagnosis/test and at-risk medical conditions for developing severe COVID-19 based on scientific evidence available on the US Centers for Disease Control and Prevention (CDC website, July 2020) and National Health Services (NHS website, July 2020) websites (see supplementary table 1).

## Data management

This study was conducted in a distributed and fully transparent manner using a publicly available common protocol[21], the publicly available ConcePTION common data model v2.2 [53] and a publicly available common analytics R-script[54]. Each data access provider (DAP) conducted the Extract-Transform-Load (ETL) process which includes a syntactic harmonization from their native data into the ConcePTION common data model (CDM). Once the data were in the CDM, data quality checks were conducted using standardized R-scripts level 1 (consistency of ETL)[55] and level 2 (logical check of data)[56]. After approval of these checks, the study analytical script could be run. Databanks available to a DAP are not always updated with the same frequency (e.g. in BIFAP not all regions link to hospital data in the same frequency). To manage with this we created subpopulations, for which follow-up time (denominator) was consistent with the period that the event could be observed.

Analysis scripts for transformation of data from the CDM into vaccines use, coverage and incidence rates were coded in R using version ≥ 3.1.0 on GitHub, where they could be downloaded for local deployment. The script is publicly available. Aggregated results of the analysis scripts were uploaded by each DAP on the Digital Research Environment (DRE) at the University Medical Center Utrecht for pooling and final analysis (www.mydre.org).

## Data analysis

Demographic characteristics including age, person-time of follow-up per age and sex group, and prevalence of underlying conditions at study entry or vaccination were computed in each data source at 1/1/2020 and at first covid-19 vaccination. For every data source, summary tables with number of administered doses per vaccine brand within the primary series (dose 1 and dose 2) by calendar time (in weeks) over the follow-up period and age at vaccination (in weeks) were created. Vaccination coverage was calculated for dose 1 and 1+2 over time. The coverage at week i was calculated by dividing the number of vaccinated subjects n_ij by the total number of subjects under follow-up at week i (N_ij), expressed as a percentage.

In each data source, crude and direct age-standardised incidence rates of each AESI (per 100,000 PYs, against the Eurostat population) were calculated both in the unvaccinated population in 2020 (background rates), and in the vaccinated cohorts, per vaccine and dose, using person time within 28 days after dose 1 and 2. For each 2-dose vaccine, we conducted analyses for each of three types of 28-day risk interval: the 28 days following Dose 1, the 28 days following Dose 2, and the days that are summed in the 28 days after either dose (total of maximum of 56 days). For each of these risk intervals, the comparator was the background rate in non-vaccinated persons in 2020. The standard population was the European Standard Population, 2013 Edition, reshaped to fit our age bands. Confidence intervals for the direct standardised rates were estimated using the formula from Fay and Feuer[57]. In addition, we calculated the difference between the standardised incidence rate post-vaccination with the background rates. The computation for the standardised incidence rates and their differences were conducted in R version 4.0.5 using the R package dsr version 0.2.2^1^.

To adjust for factors associated with exposure to covid-19 vaccines we used a multivariate Poisson regression adjusting concurrently for the four factors that were included in the original monitoring protocol (age, gender, any risk factor for covid-19 disease severity, and previous Sars-Cov-2 infection). The log person-days in each risk or comparison interval were included in the regression model as the offset. Incidence rate ratios – estimating the ratio of outcome incidence in the risk interval divided by outcome incidence in the comparison interval are reported with 95% confidence intervals. Specific analyses were conducted for each brand and the dose 1 and 2 risk interval of covid-19 vaccine against the non-exposed. The negative binomial regression model did not converge thus exact Poisson interval were calculated. Since the number of events were low, incidence rate ratios across DAPs were pooled using a random effects model using the R package *meta*, this was conducted as a post-hoc analysis. Effect modification could not be explored due to limited number of cases in each of the data sources for the very rarely occurring AESI.

## Results

This study comprised a total of 25,720,158 subjects (table 1). We count only the largest population for BIFAP for the total, as the regions with hospital linkage are a subset of the primary care populations. The largest population included was from CPRD followed by BIFAP. Data locks differed per site, the recommended end date to use the data was June 30, 2021 in Tuscany, August 31st for BIFAP, August 1st 2021 for PHARMO and May 2021 for CPRD Aurum (table 1).

Table 2 shows the characteristics of the study population at the start of the study. Median age at study start was highest in Tuscany (49 years) and in the region of BIFAP that could be linked to hospitalizations, median age was lowest in the population covered in the PHARMO data source (40 years). Prevalence of any risk factor for severe covid-19 was highest in Tuscany (34.4%) whereas it was around 25% in each of the other data sources (table 2).

**Table 2.**
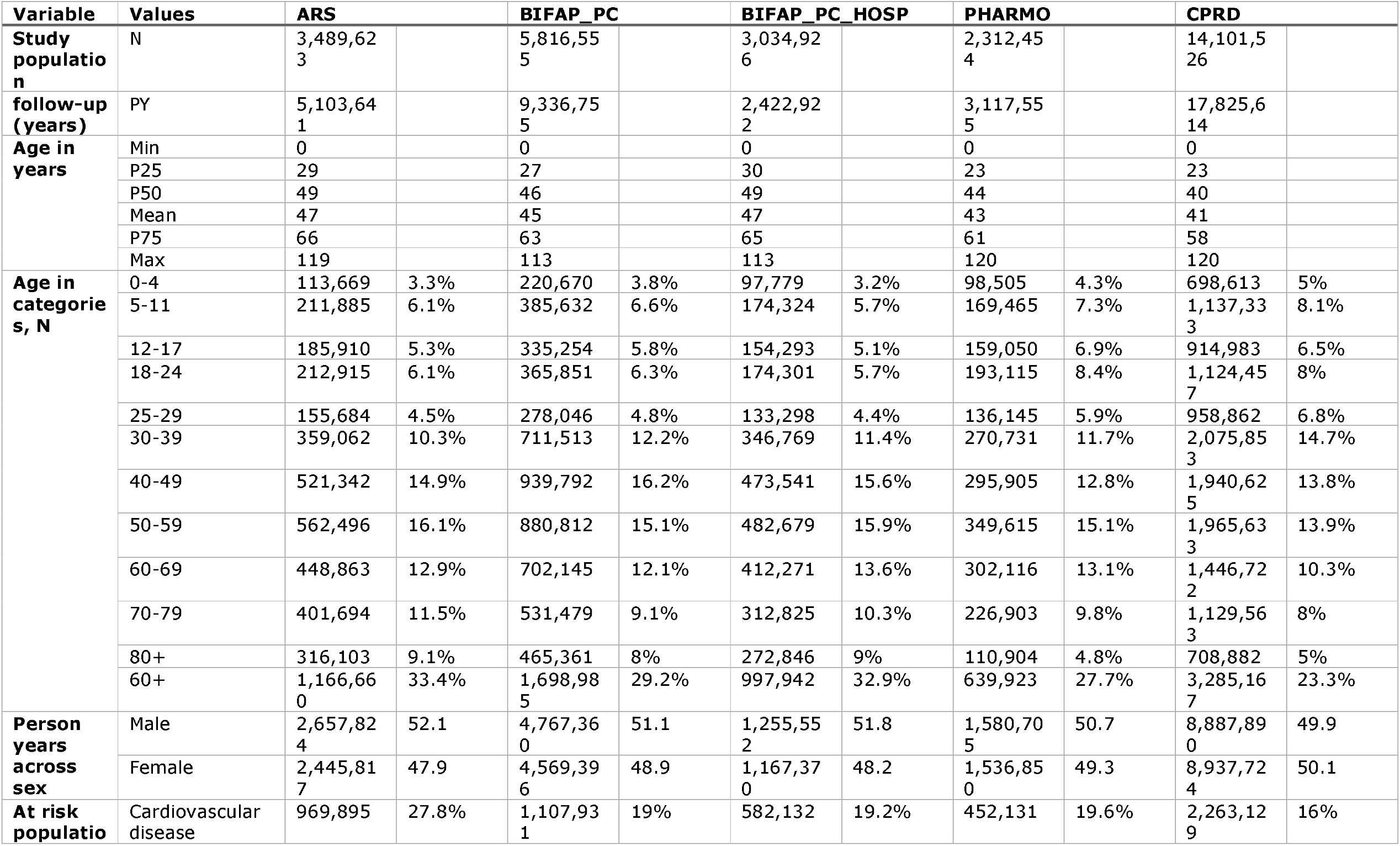
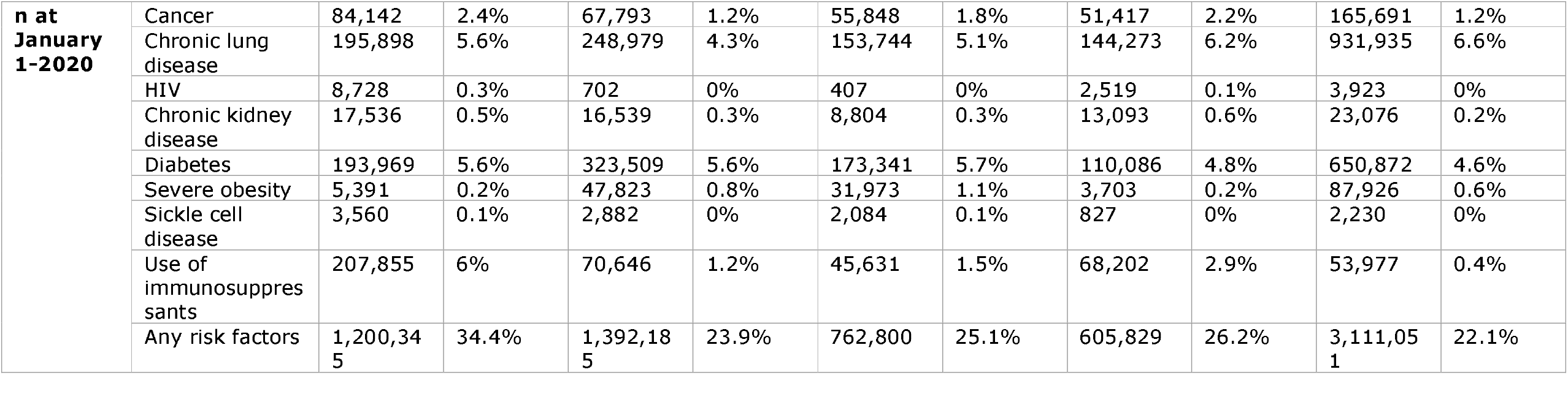
Baseline characteristics of the study population for the cohort study at 1/1/2020.

Table 3 shows how the different countries used different vaccination strategies until the data lock point. In the study population for this project 12,117,458 persons received at least a first dose of covid-19 vaccine: 54% with Comirnaty, 6% Spikevax, 38% Vaxzevria and 2% Janssen COVID-19 vaccine, across all data sources (counting only BIFAP-primary care).ARS and CPRD could capture all the brands, only PHARMO and BIFAP had covid-19 vaccines with unknown brands (233 doses for BIFAP and 113,201 doses for PHARMO, see tables S3 and S4). In the data instance for this analysis, a varying percentage of persons with a first dose of a specific covid-19 vaccine had received a second dose, this was highest in BIFAP and Tuscany. The distance between first and second dose was longest in the CPRD, especially for Comirnaty (median 77 days). In ARS, BIFAP and the PHARMO data sources, Comirnaty and Spikevax second doses were at shorter distance in those with a second recorded dose (within approximately 21-35 days after each other), but the distance of Vaxzevria doses was similar to the CPRD. Very few people received heterologous schemes (a different brand of first and second dose) in this data instance (table 3).

**Table 3:** Vaccinations and distances between doses across COVID-19 vaccines.

| Dose | Measure | ARS |  | BIFAP_<br>PC |  | BIFAP_<br>PC_HO<br>SP |  | PHARM<br>O |  | CPRD |  | Total |  |
| --- | --- | --- | --- | --- | --- | --- | --- | --- | --- | --- | --- | --- | --- |
| Study population | N | 348962<br>3 |  | 581655<br>5 |  | 303492<br>6 |  | 231245<br>4 |  | 141015<br>26 |  | 257201<br>58 |  |
| AstraZeneca dose 1, % of total population | Persons | 332872 | 9.5% | 537122 | 9.2% | 78602 | 2.6% | 68655 | 3.0% | 367167<br>2 | 26.0% | 461032<br>1 | 17.9% |
| AstraZeneca dose 2, % of 1st dose | Persons | 187052 | 56% | 397186 | 74% | 59342 | 75% | 28779 | 42% | 117274<br>5 | 32% | 178576<br>2 | 39% |
| Other vaccine dose 2, % of first dose | Persons | 7150 | 2% | 7298 | 1% | 368 | 0% | 0 | 0% | 3113 | 0% | 17561 | 0% |
| Amongst persons with AstraZeneca dose 2 distance | Min | 20 |  | 14 |  | 21 |  | 70 |  | 14 |  |  |  |
| Amongst persons with AstraZeneca dose 2 distance | P25 | 84 |  | 71 |  | 70 |  | 76 |  | 70 |  |  |  |
| Amongst persons with AstraZeneca dose 2 distance | P50 | 84 |  | 82 |  | 76 |  | 77 |  | 77 |  |  |  |
| Amongst persons with AstraZeneca dose 2 distance | P75 | 84 |  | 84 |  | 84 |  | 84 |  | 78 |  |  |  |
| Amongst persons with AstraZeneca dose 2 distance | Max | 126 |  | 193 |  | 166 |  | 155 |  | 127 |  |  |  |
| Janssen dose 1, % of total population | Persons | 58513 | 1.7% | 201543 | 3.5% | 31993 | 1.1% | 22455 | 1.0% |  |  | 282511 | 1.1% |
| Janssen dose 2, % of 1st dose | Persons | 0 |  | 0 |  | 0 |  | 0 |  |  |  | 0 |  |
| Other vaccine dose 2. % of 1st dose | Persons | 0 |  | 63 | 0.0% | 11 | 0.0% | 15 | 0.1% |  |  | 78 | 0.0% |
| Moderna dose 1, % of total population | Persons | 184013 | 5.3% | 447401 | 7.7% | 74275 | 2.4% | 67689 | 2.9% | 27023 | 0.2% | 726126 | 2.8% |
| Moderna dose 2, % of 1st dose | Persons | 100673 | 54.7% | 363226 | 81.2% | 60459 | 81.4% | 25638 | 37.9% | <5 |  | 489540 | 67.4% |
| Other vaccine dose 2, % of 1st dose | Persons | 125 | 0.1% | 590 | 0.1% | 37 | 0.0% | 0 | 0.0% | 9 | 0.0% | 718 | 0.1% |
| Amongst persons with Moderna dose 2 distance | Min | 16 |  | 14 |  | 14 |  | 21 |  | 28 |  |  |  |
| Amongst persons with Moderna dose 2 distance | P25 | 28 |  | 28 |  | 28 |  | 35 |  | 28 |  |  |  |
| Amongst persons with Moderna dose 2 distance | P50 | 28 |  | 28 |  | 28 |  | 35 |  | 28 |  |  |  |
| Amongst persons with Moderna dose 2 distance | P75 | 28 |  | 28 |  | 28 |  | 35 |  | 44 |  |  |  |
| Amongst persons with Moderna dose 2 distance | Max | 124 |  | 224 |  | 224 |  | 160 |  | 91 |  |  |  |
| Pfizer dose 1. % of total population | Persons | 1320326 | 37.8% | 2808700 | 48.3% | 353509 | 11.6% | 568119 | 24.6% | 1801355 | 12.8% | 6498500 | 25.3% |
| Pfizer dose 2. % of 1st dose | Persons | 653580 | 49.5% | 2372395 | 84.5% | 308848 | 87.4% | 232351 | 40.9% | 1332285 | 74.0% | 4590611 | 70.6% |
| Other vaccine dose 2, % of 1st dose | Persons | 138 | 0.0% | 1179 | 0.0% | 30 | 0.0% | 0 | 0.0% | 6226 | 0.3% | 7543 | 0.2% |
| Amongst persons with Pfizer dose 2 distance | Min | 14 |  | 14 |  | 14 |  | 21 |  | 14 |  |  |  |
| Amongst persons with Pfizer dose 2 distance | P25 | 21 |  | 21 |  | 21 |  | 35 |  | 70 |  |  |  |
| Amongst persons with Pfizer dose 2 distance | P50 | 21 |  | 21 |  | 21 |  | 35 |  | 76 |  |  |  |
| Amongst persons with Pfizer dose 2 distance | P75 | 21 |  | 21 |  | 21 |  | 36 |  | 78 |  |  |  |
| Amongst persons with Pfizer dose 2 distance | Max | 174 |  | 244 |  | 210 |  | 169 |  | 147 |  |  |  |
| **Total first doses, % of total population** | **Persons** | **1895724** | **54.3%** | **3994766** | **68.7%** | **538379** | **17.7%** | **726918** | **31.4%** | **5500050** | **39.0%** | **12117458** | **47.1%** |

Characteristics of the recipients of the different type of COVID-19 vaccines recipients are provided in the supplementary materials (Tables 2-6). To summarize: In Tuscany (table S2), the data instance for this analysis was updated until June 2021, people with vaccination had high prevalence of at-risk conditions for serious COVID-19 >50% at first covid-19 vaccination for each of the vaccines. Among persons vaccinated with Comirnaty, 21.9% were 80+, which was a big difference with other covid-19 vaccines which all had less than 1% in the above 80 group. Vaxzevria and Janssen vaccine were provided primarily to persons between 50 and 79.

For Spain data was provided until August 2021 for primary care data in BIFAP (table S3), the percentage of at-risk conditions was highest for Vaxzevria (41.7%) followed by Comirnaty (38.3%) Spikevax vaccine recipients (32%) and lowest for Janssen Covid-19 vaccine (28%). Vaxzevria was provided almost exclusively to those 50-69 years of age (82.9% of Vaxzevria recipients), 80+ received mostly Comirnaty, or Spikevax.

For the Netherlands PHARMO included data until July, 2021, prevalence of at-risk conditions were highest in those vaccinated with Vaxzevria (54.1%) followed by Comirnaty (44.2%) Spikevax (28.8%) and Janssen (9.4%) (table S4). Vaxzevria recipients were almost all between 60-69 years of age (86%) in the Netherlands. Median age for those receiving Janssen Covid-19 vaccine was 26, and almost only young adults received this vaccine.

In CPRD (UK), data was available until May 2021 (table S4). Majority of persons received Vaxzevria (66.8%), characteristics of Vaxzevria users and Comirnaty recipients differed for Comirnaty: 20% of Comirnaty users was over 80 years of age, whereas this was only 3.9% for Vaxzevria. The prevalence of at-risk conditions in those receiving Comirnaty (59%) was also much higher than in those with Vaxzevria (42.1%), recipients of Spikevax had low prevalence of at-risk conditions (13.3%) and were mostly in the 40-49 years of age range (91.1%). No Janssen vaccine was administered.

Case counts and crude and age standardized incidence rates and rate differences for each of the data sources and each of the AESI for non-vaccinated in 2020 and post-COVID-19 vaccination are provided in the supplementary tables S6.

Fourteen of the AESI were very rare with pooled age standardized background incidence rates < 10/100,000 PY (table 4 for pooled data and table S6 for individual data sources data). This included (all per 100,000 PY) ADEM (1.05), acute liver injury (IR=8.45), disseminated intravascular coagulation (0.27), erythema multiforme (4.81), GBS (1.74), meningoencephalitis (4.4), microangiopathy (0.73), multi-inflammatory syndrome (0.83), myocarditis (4.1), narcolepsy (1.0), single organ cutaneous vasculitis (5.4), stress cardiomyopathy (2.0), transvers myelitis (1.0) and thrombotic thrombocytopenia syndrome (0.56). AESI with incidence rates of rare events between 10 and 100/100,000 PY comprised seven conditions: anaphylaxis (11.7), anosmia/ageusia (71.3), acute respiratory distress syndrome (37.9), Bell’s palsy (29.1), Chilblain like lesions (17.1), hemorrhagic stroke (25.1/100,000), myo/pericarditis (14.7) thrombocytopenia (27.0). All other conditions were more common with incidence rates (>100 /100,000) which included acute kidney injury (132), acute coronary artery disease (129), arrhythmia (598), heart failure (209), ischemic stroke (117), venous thromboembolism (130/100,000) varied between countries, as well as generalized convulsions (135/100,000). Death was common with 720/100,000PY. Incidence rates were quite similar across data sources, largest variations were observed between data sources with different provenance of diagnoses (e.g. including hospital (ARS, BIFAP-PC-HOSP), emergency room (ARS), or only primary care (BIFAP-PC, PHARMO, CPRD) for conditions that are not typically diagnosed in hospital such as chilblains, anosmia/ageusia and anaphylaxis (mostly emergency room in ARS). Table 4 and S6 shows the age standardized incidence rate differences for pooled data (Table 4) and individual data sources (S6). Age standardized pooled rate differences across data sources were statistically significantly elevated for acute kidney injury following Vaxzevria dose 1 and 1&2, anaphylaxis after Vaxzevria dose 1 and 1&2, ARDS after Spikevax dose 1 and dose 1&2, arrhythmia after Spikevax dose 1, 2 and 1&2, chilblain like lesions after Vaxzevria dose 1 and dose 1&2 and Comirnaty dose 1 and 1&2, death after Vaxzevria dose 1 and dose 1&2, generalized convulsions after Vaxzevria dose 1,2 and dose 1&2, heart failure after Vaxzevria dose 1 and dose 1&2 as well as Spikevax dose 1,2 and dose 1&2, and Comirnaty dose 1, 2 and dose 1&2, ischemic stroke after Vaxzevria dose 1&2, myo/pericarditis after Comirnaty dose 2 and dose 1&2, thrombocytopenia after dose 1, 2 1&2 of Vaxzevria, Spikevax dose 1&2, and Comirnaty dose 1, 2 and 1&2, VTE after Vaxzevria dose 1, 1&2, Spikevax 1, 2, and 1&2, Comirnaty 1, 2 and 1&2, TTS after Vaxzevria dose 1 and 1&2. After adjustment for the factors associated with vaccination exposure using a Poisson regression, ten pooled (random effects) associations remained for dose 1&2 28-day risk intervals combined, these included anaphylaxis after Vaxzevria (IRR=1.68, 95%CI 1.37-2.1), erythema multiforme after Spikevax (IRR=2.64, 95%CI 1.25-5.60), GBS after Janssen dose 1 (IRR=5.7, 95%CI: 1.4-23), SOCV after Janssen dose 1 (IRR=4.4, 95%CI 1.1-17.7), thrombocytopenia after Janssen dose 1 (IRR=2.3, 95%CI 1.3-4.1), Spikevax (IRR=1.8, 95%CI: 1.1-3.2), VTE after Spikevax (IRR=1.6, 95%CI 1.4-1.8) and Comirnaty (IRR=1.1, 95%CI 1.0-1.2), TTS after Vaxzevria (IRR=2.98, 95%CI: 1.67-5.31) and after Janssen dose 1 (IR=90,10-infinity), only 5 combinations had IRR above 2.

**Table 4:**
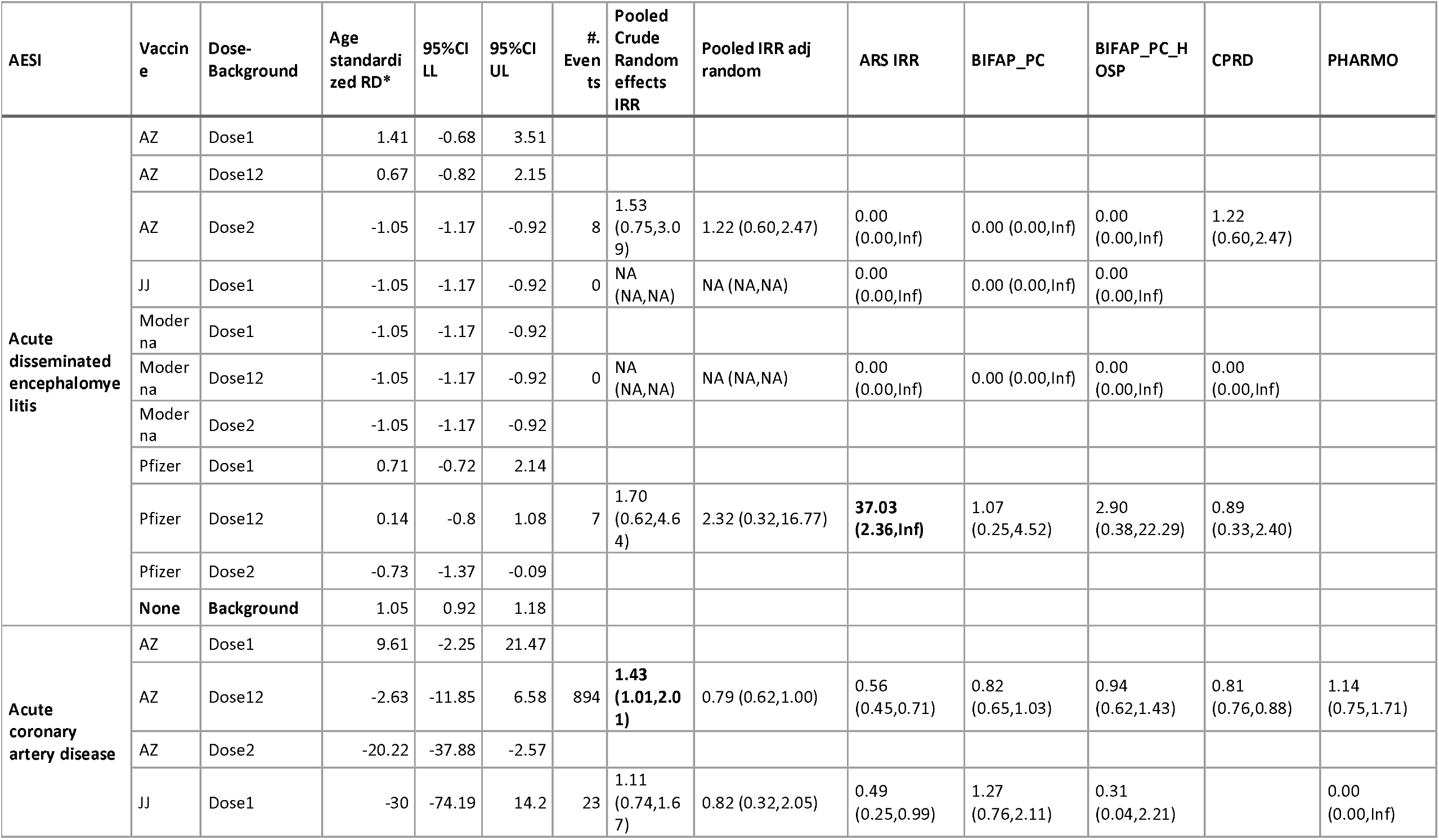
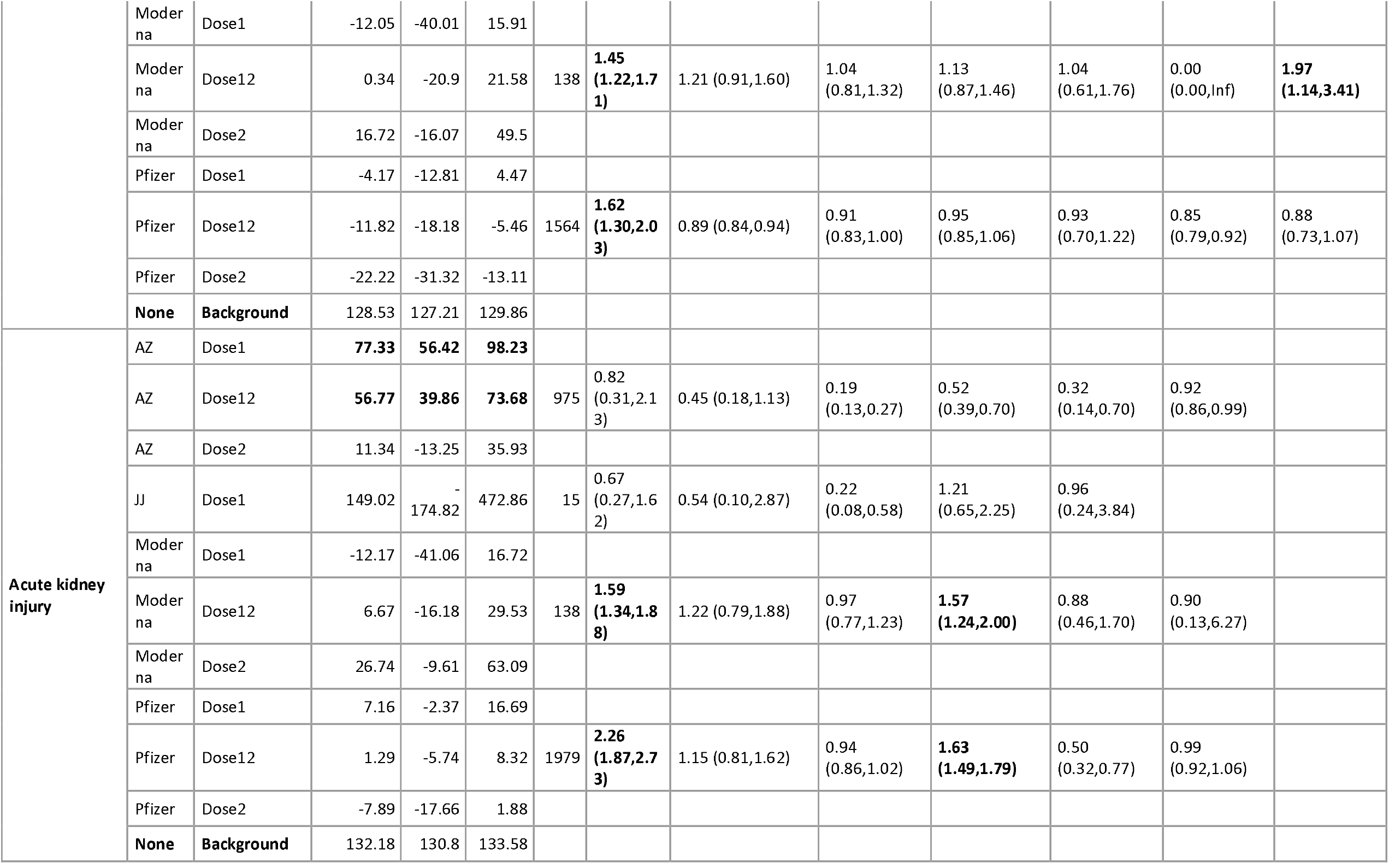
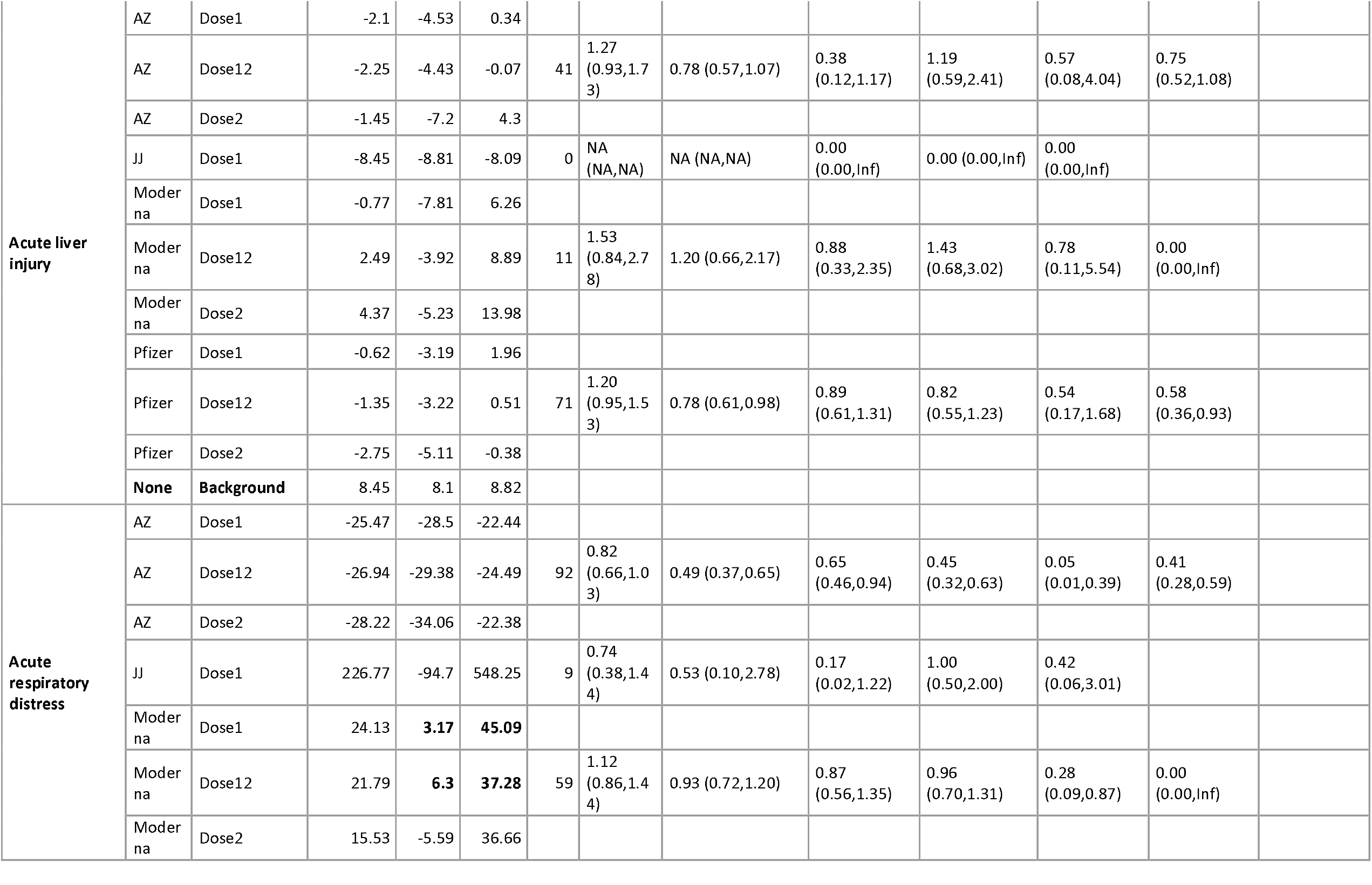
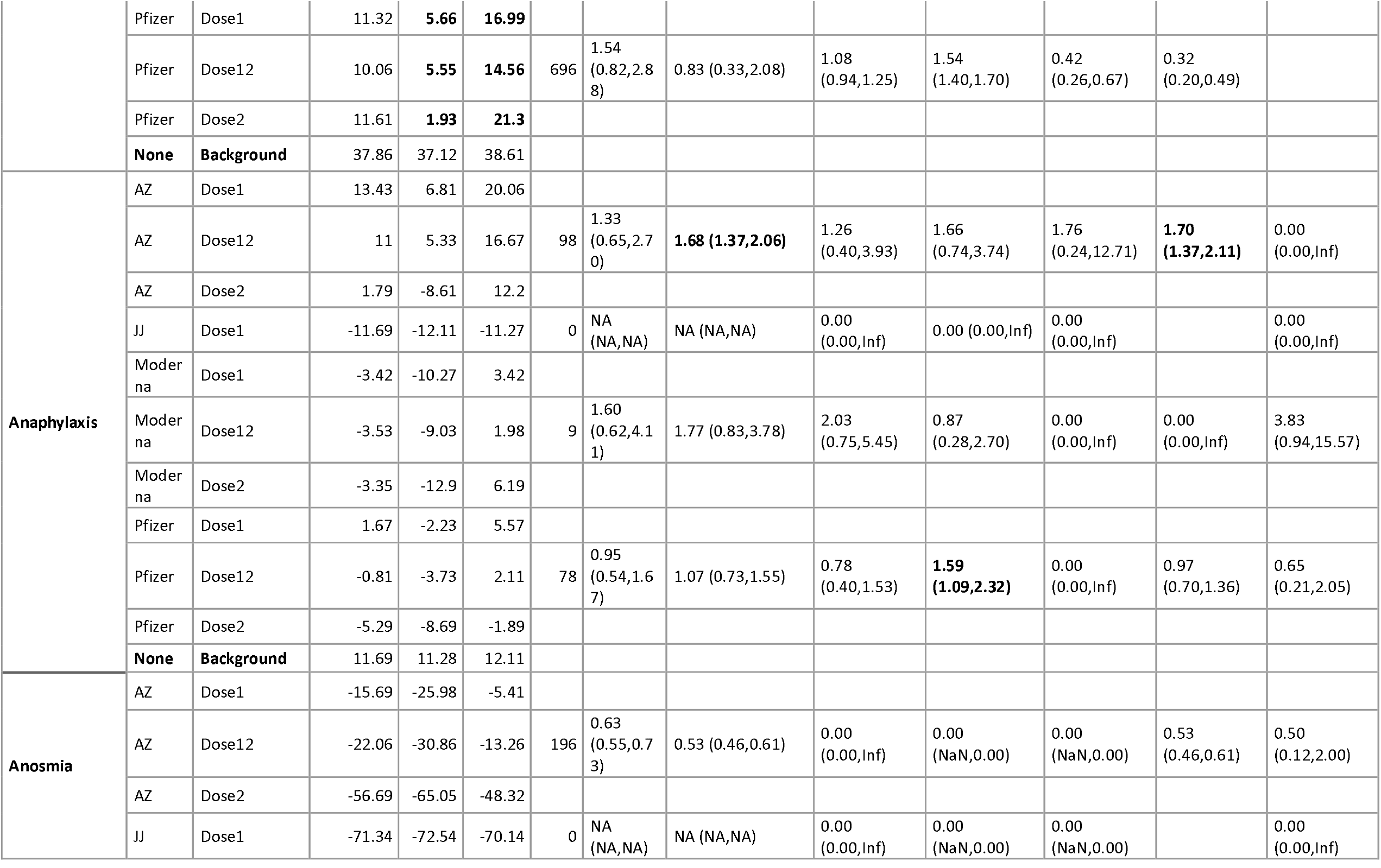
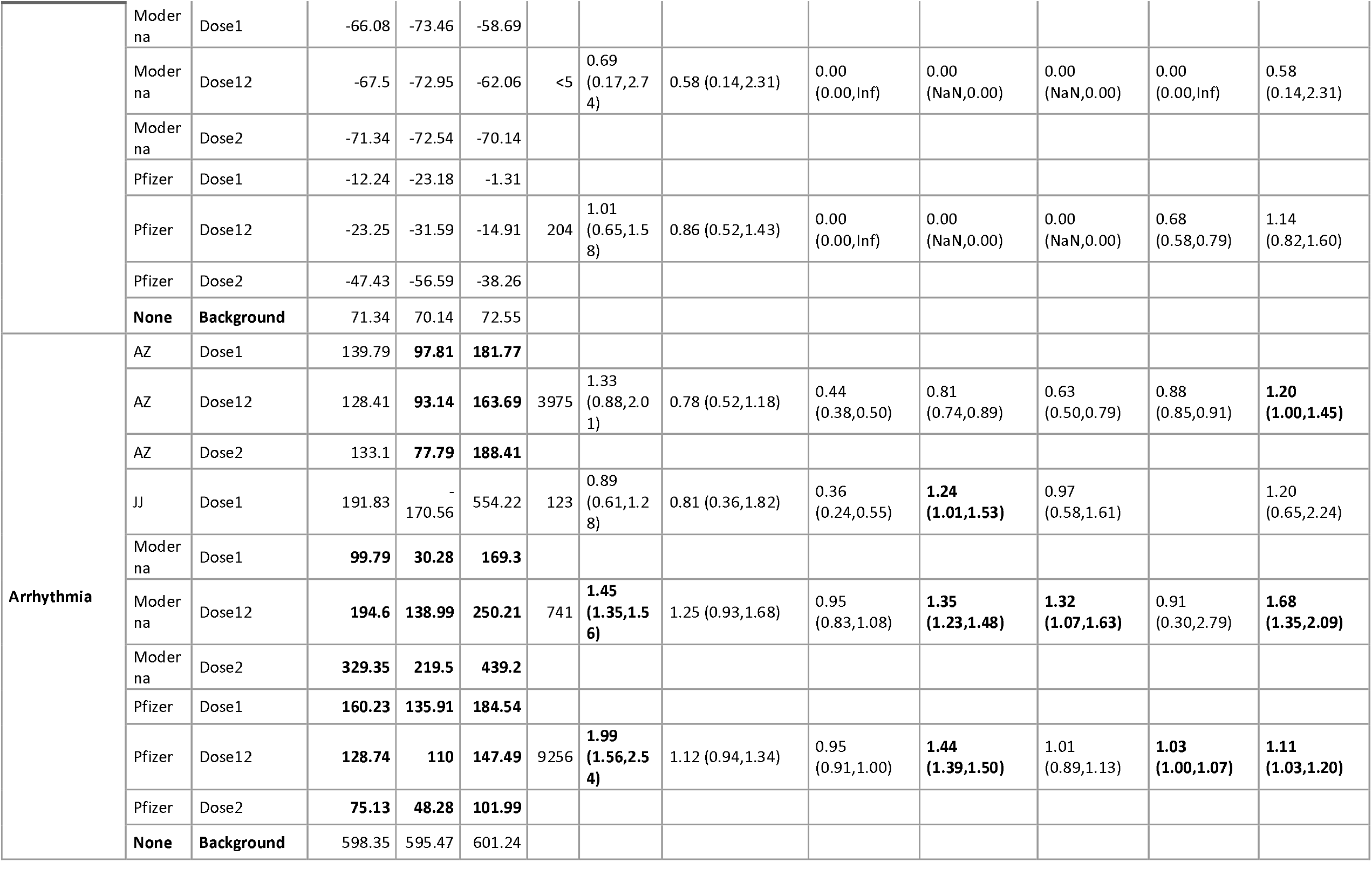
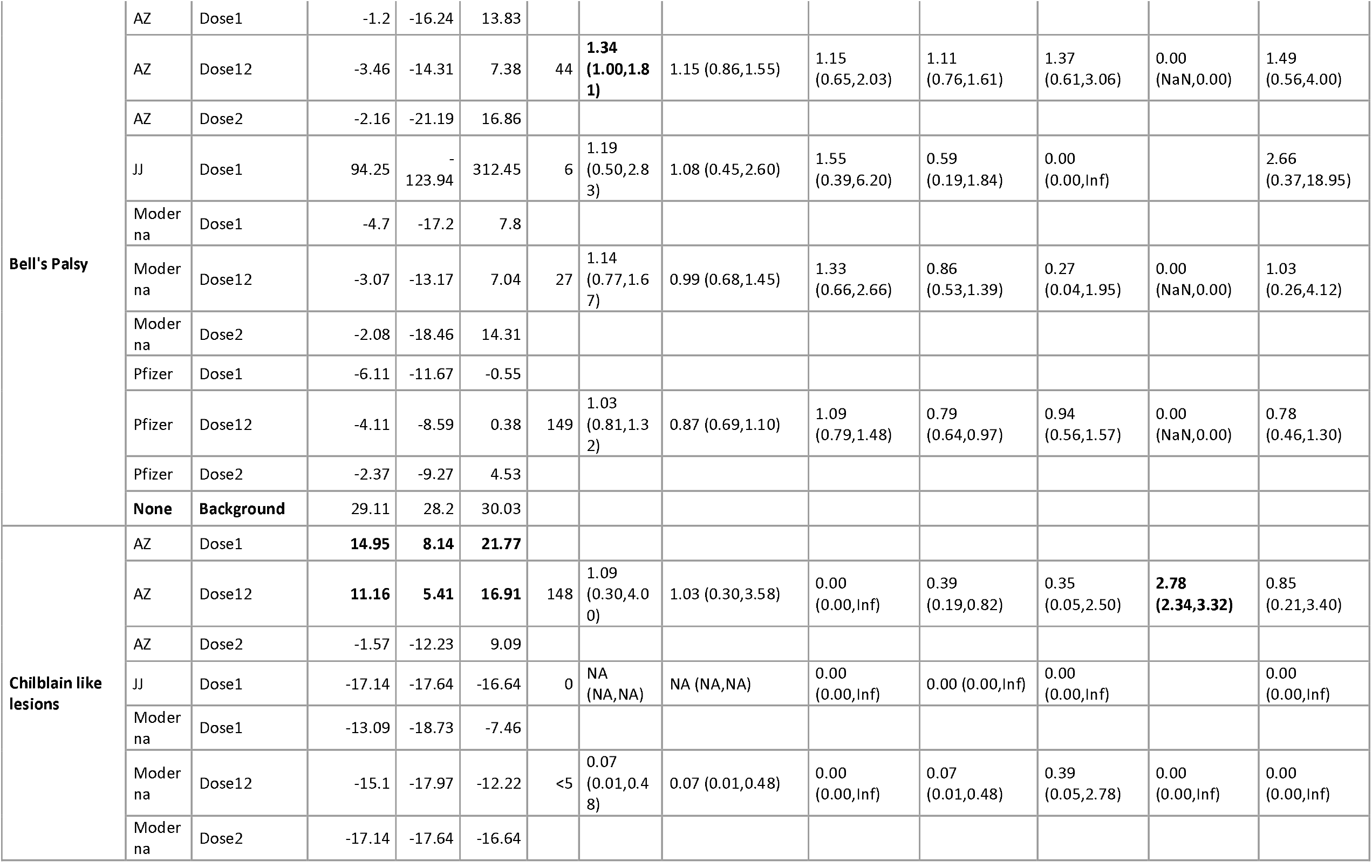
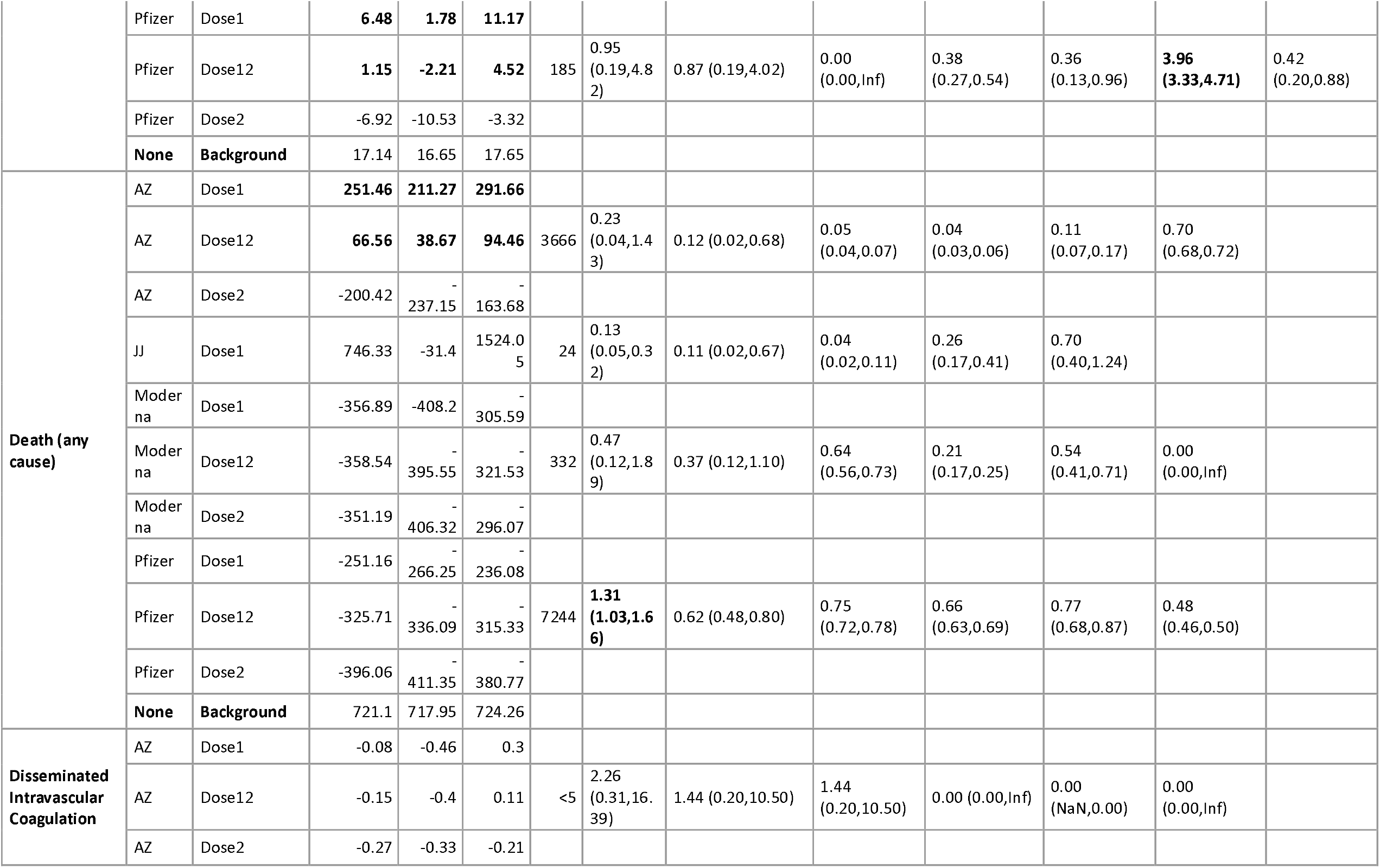
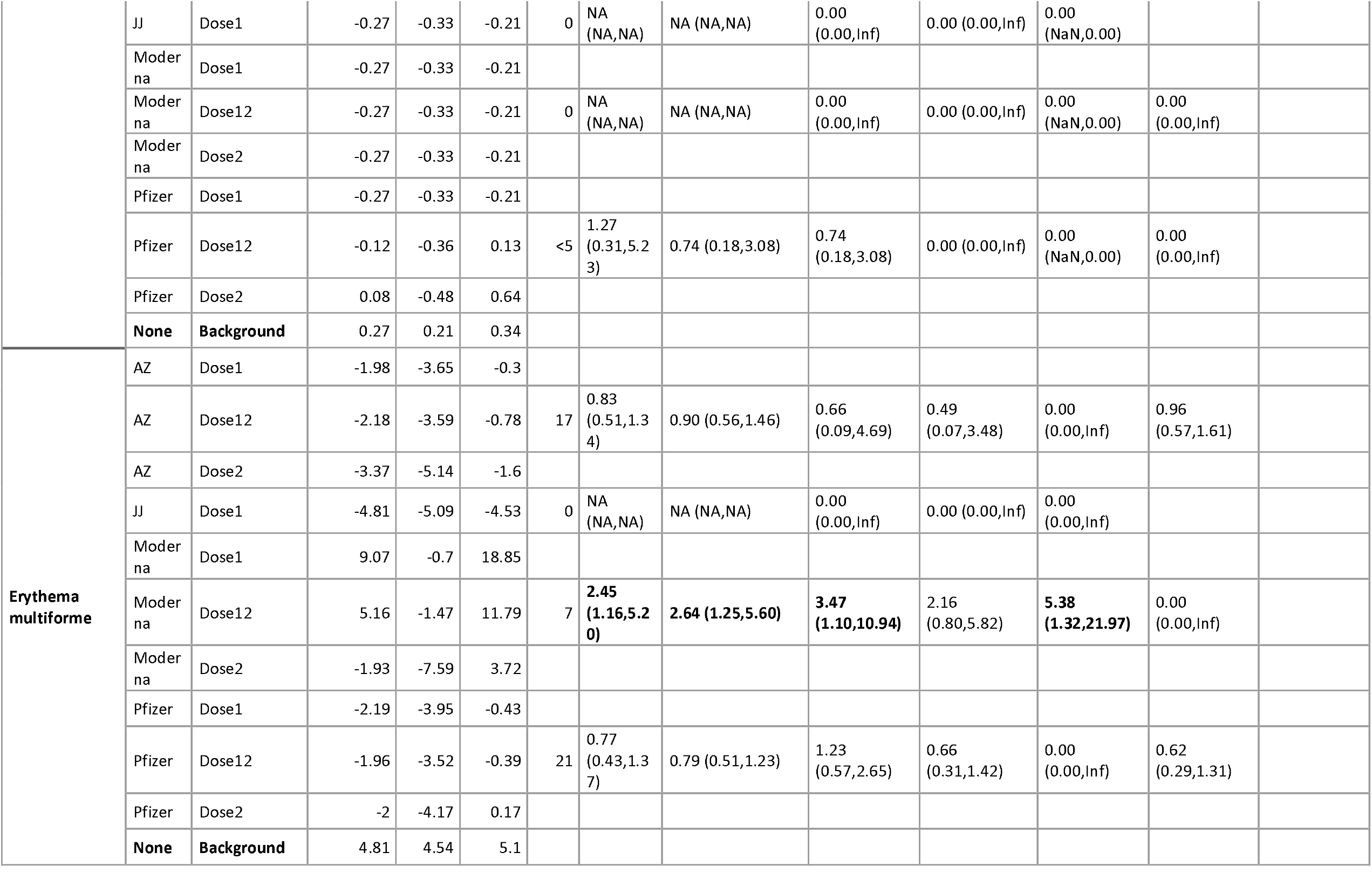
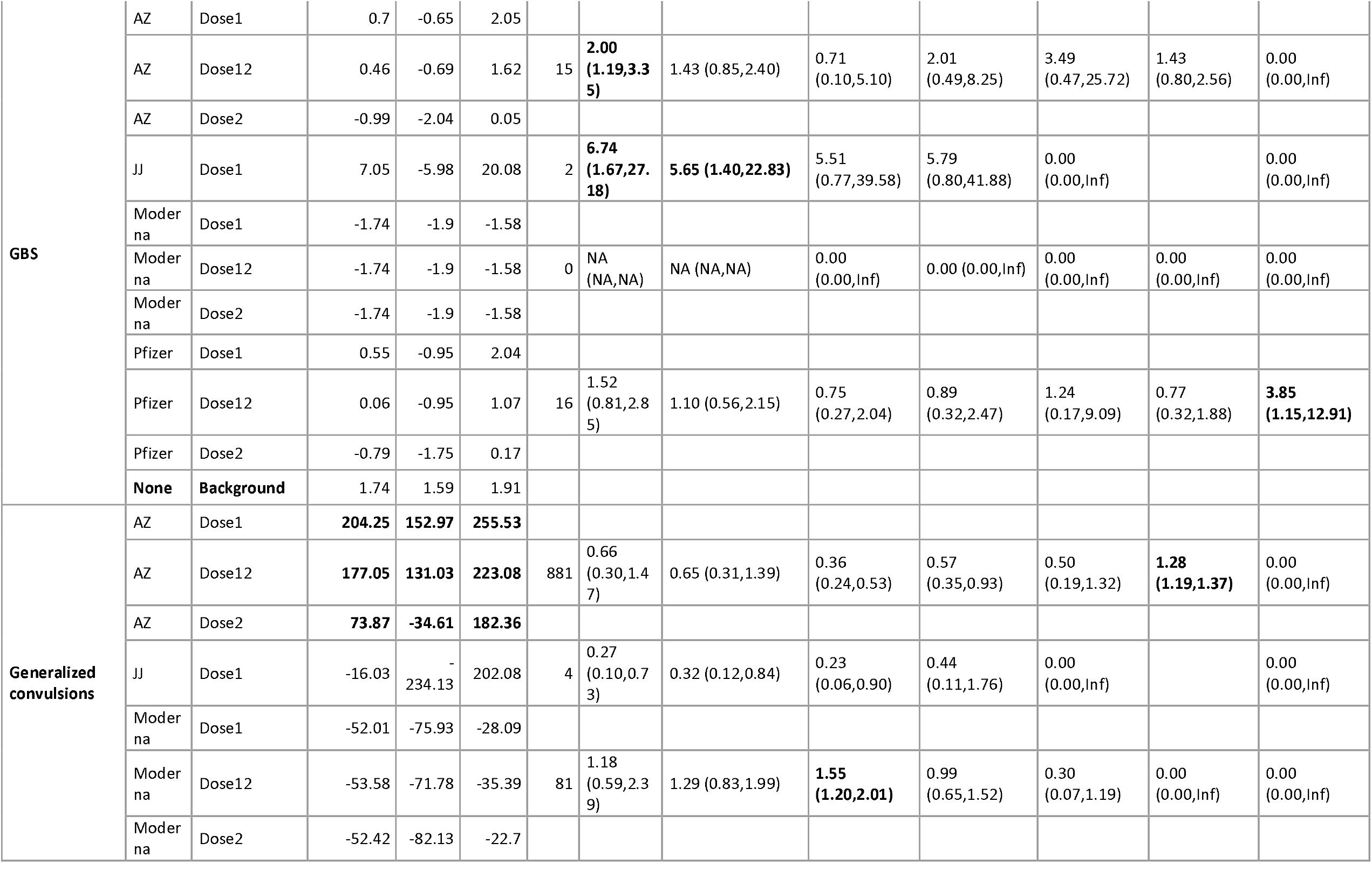
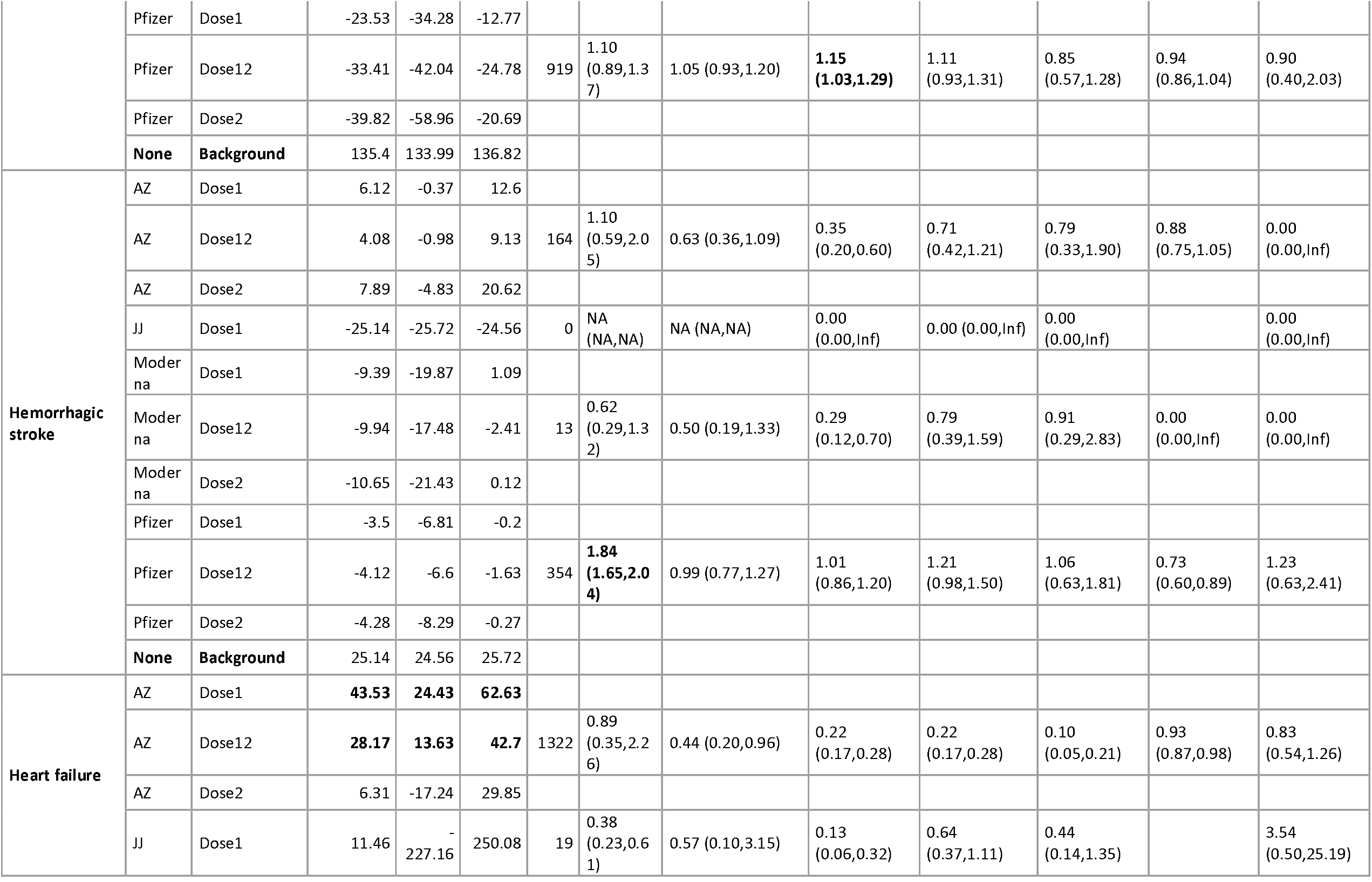
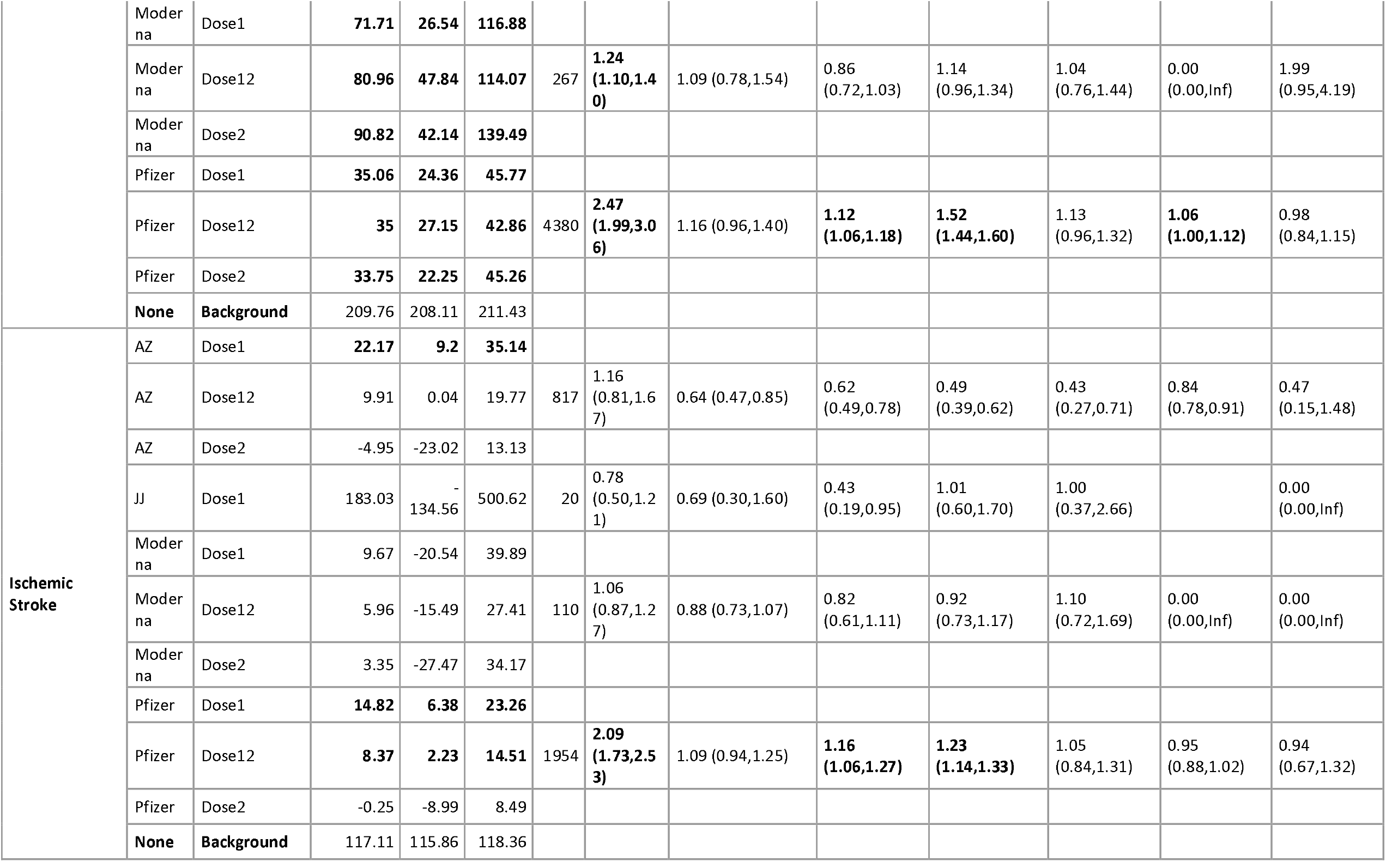
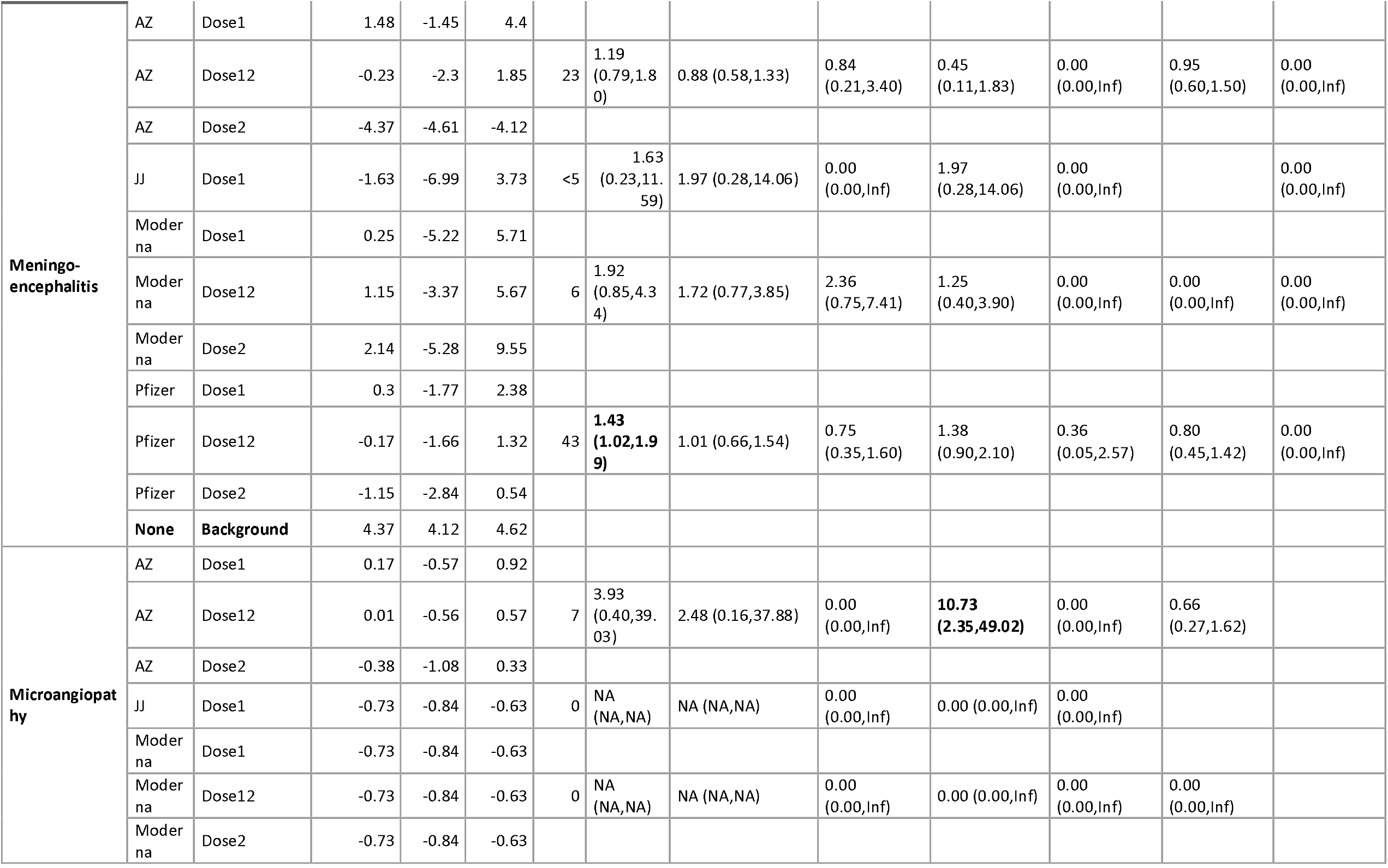
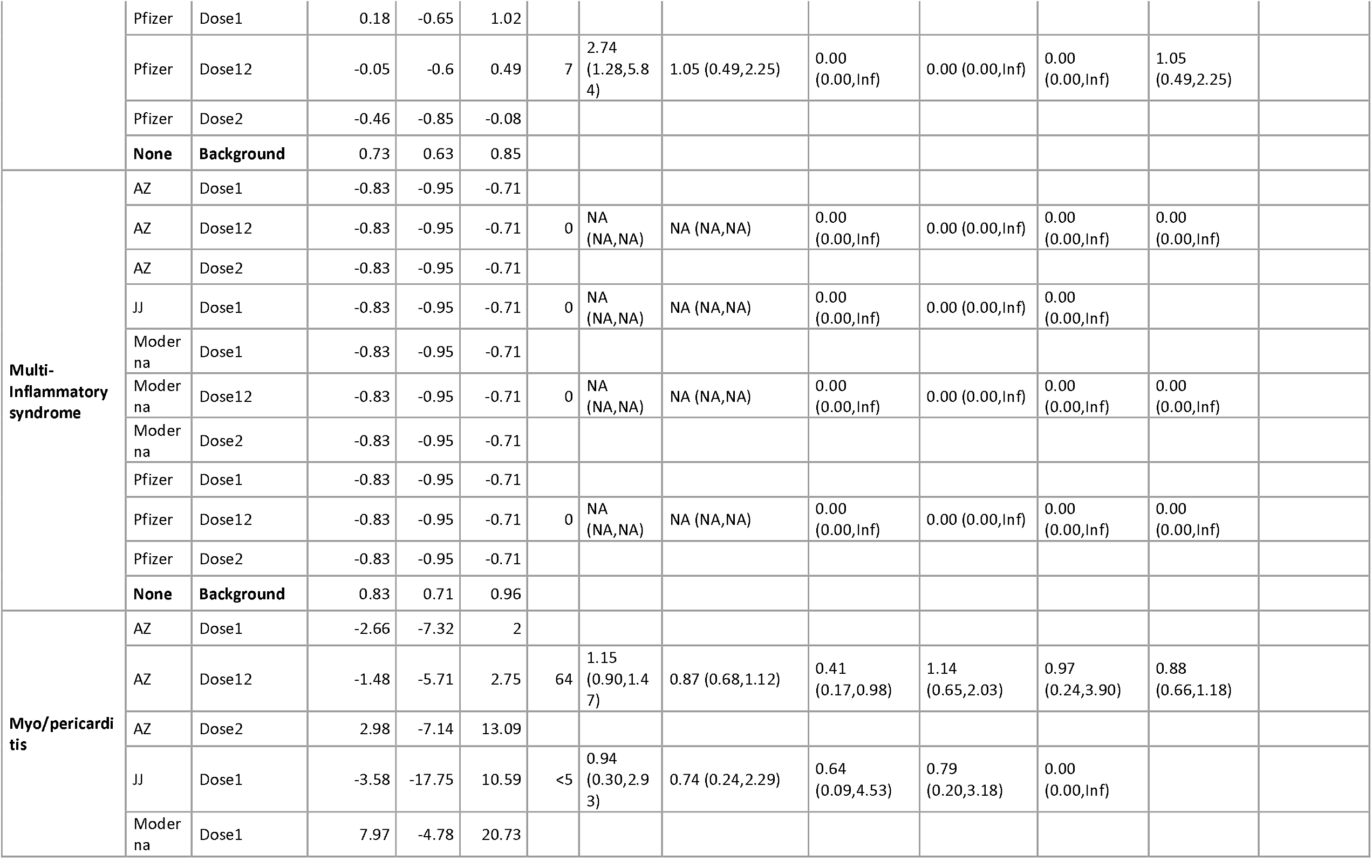
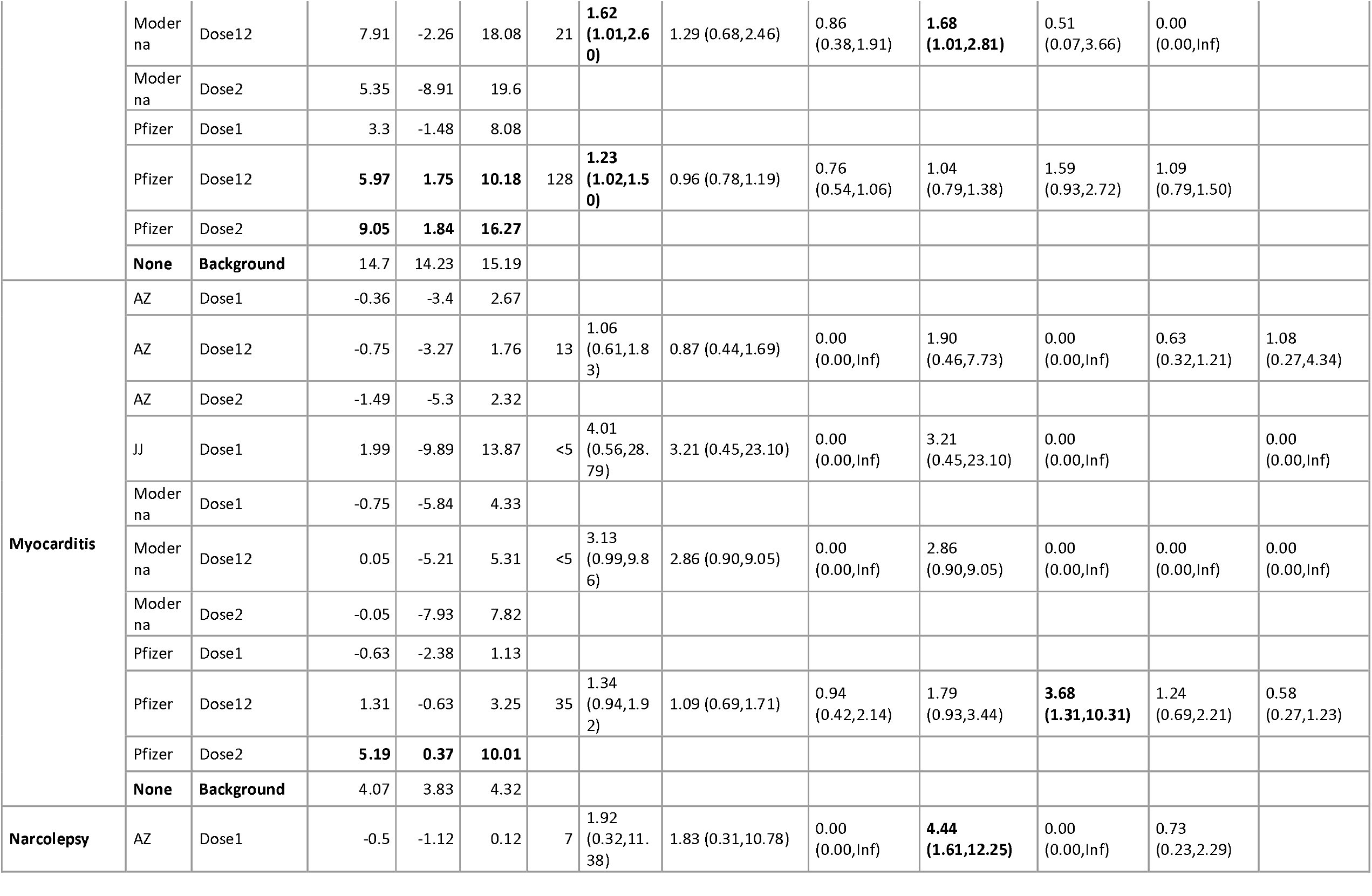
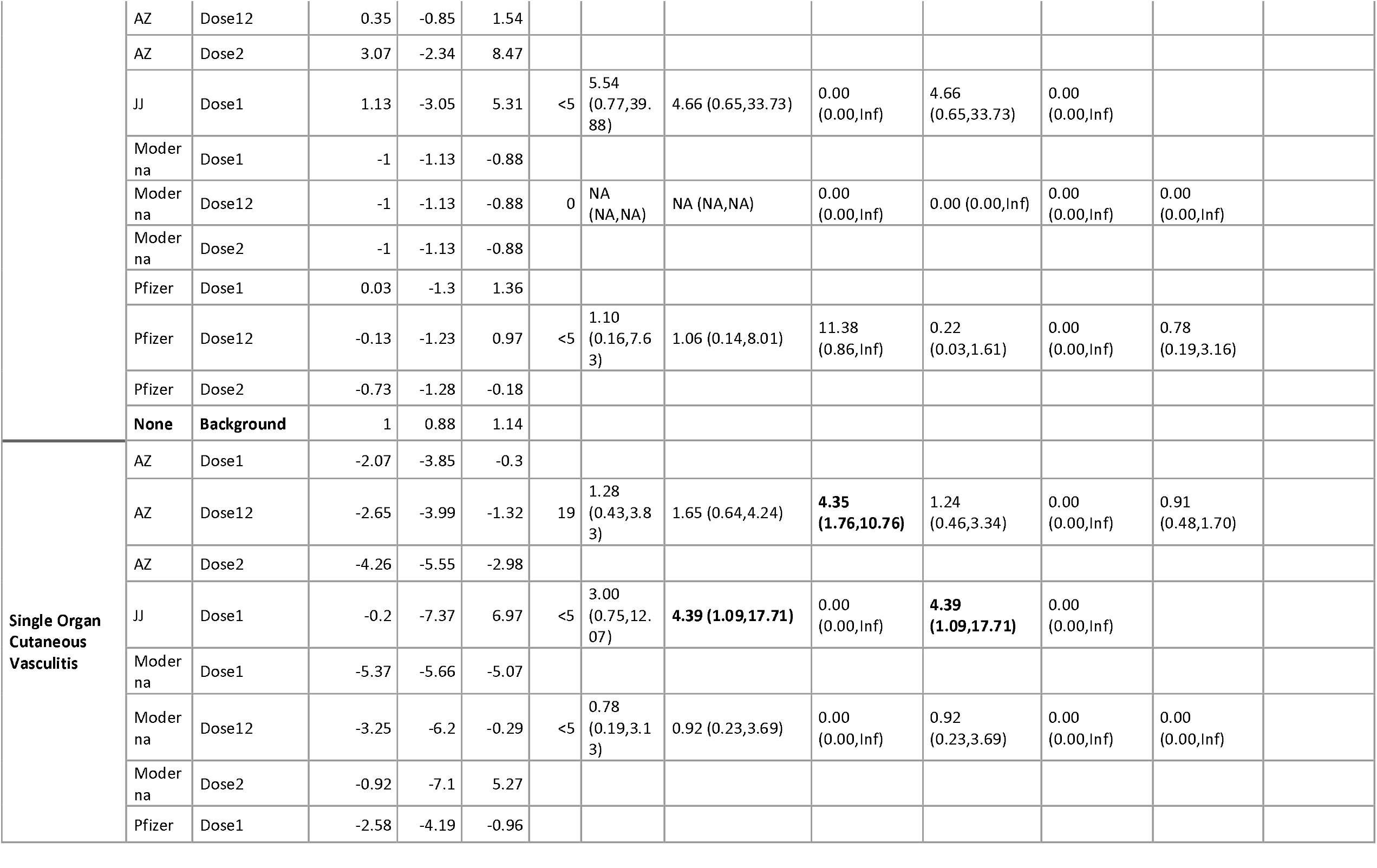
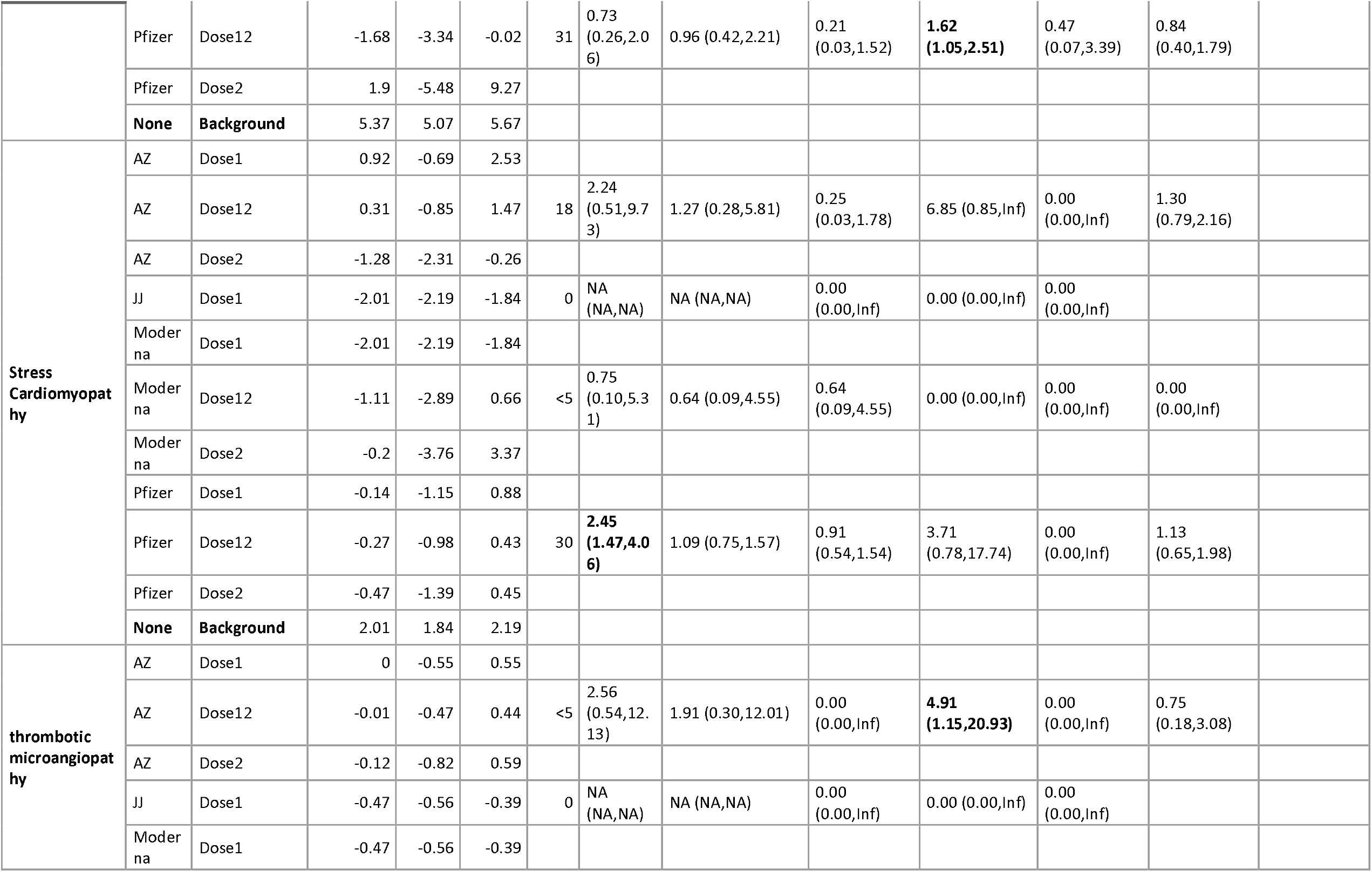
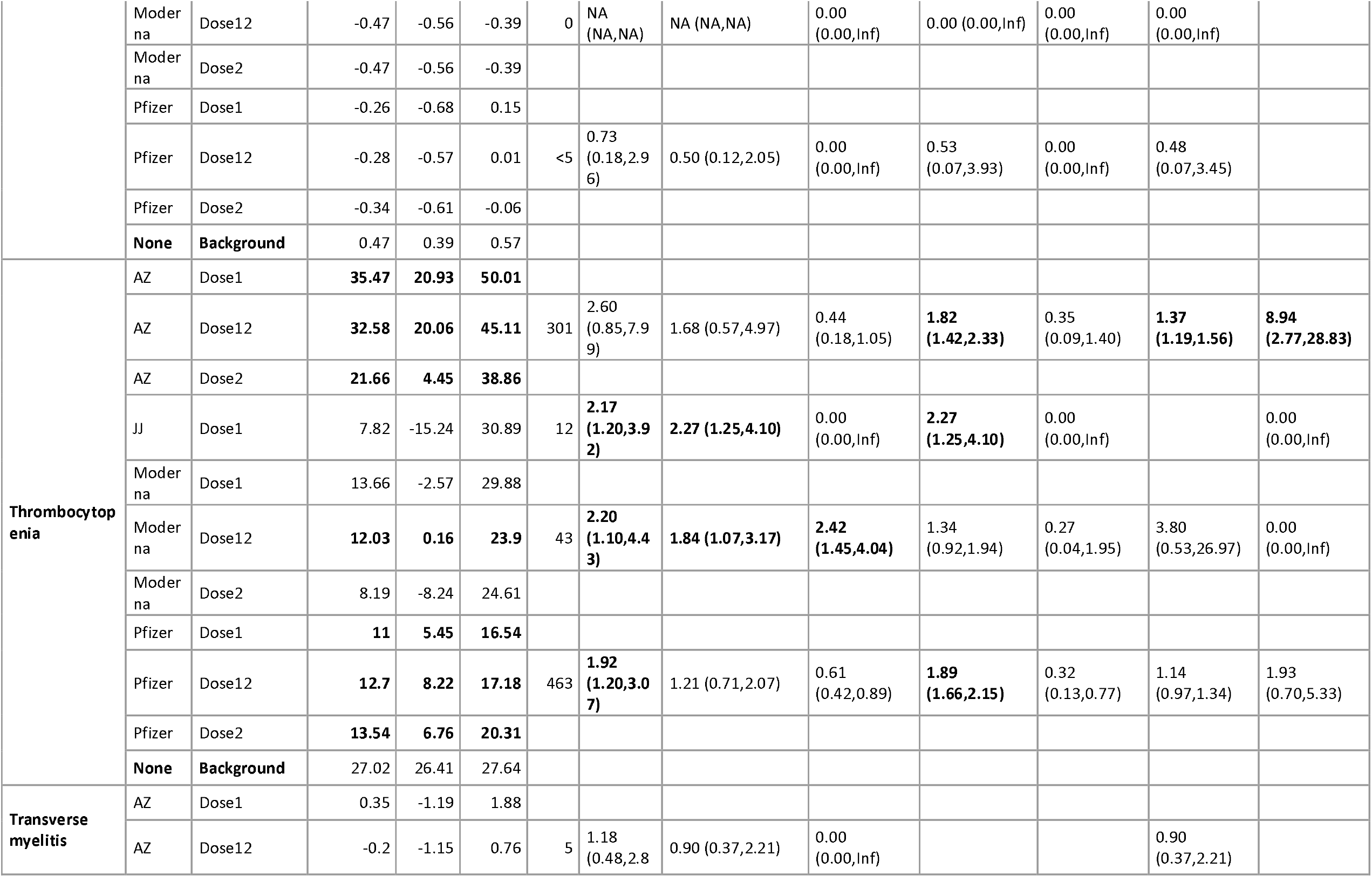
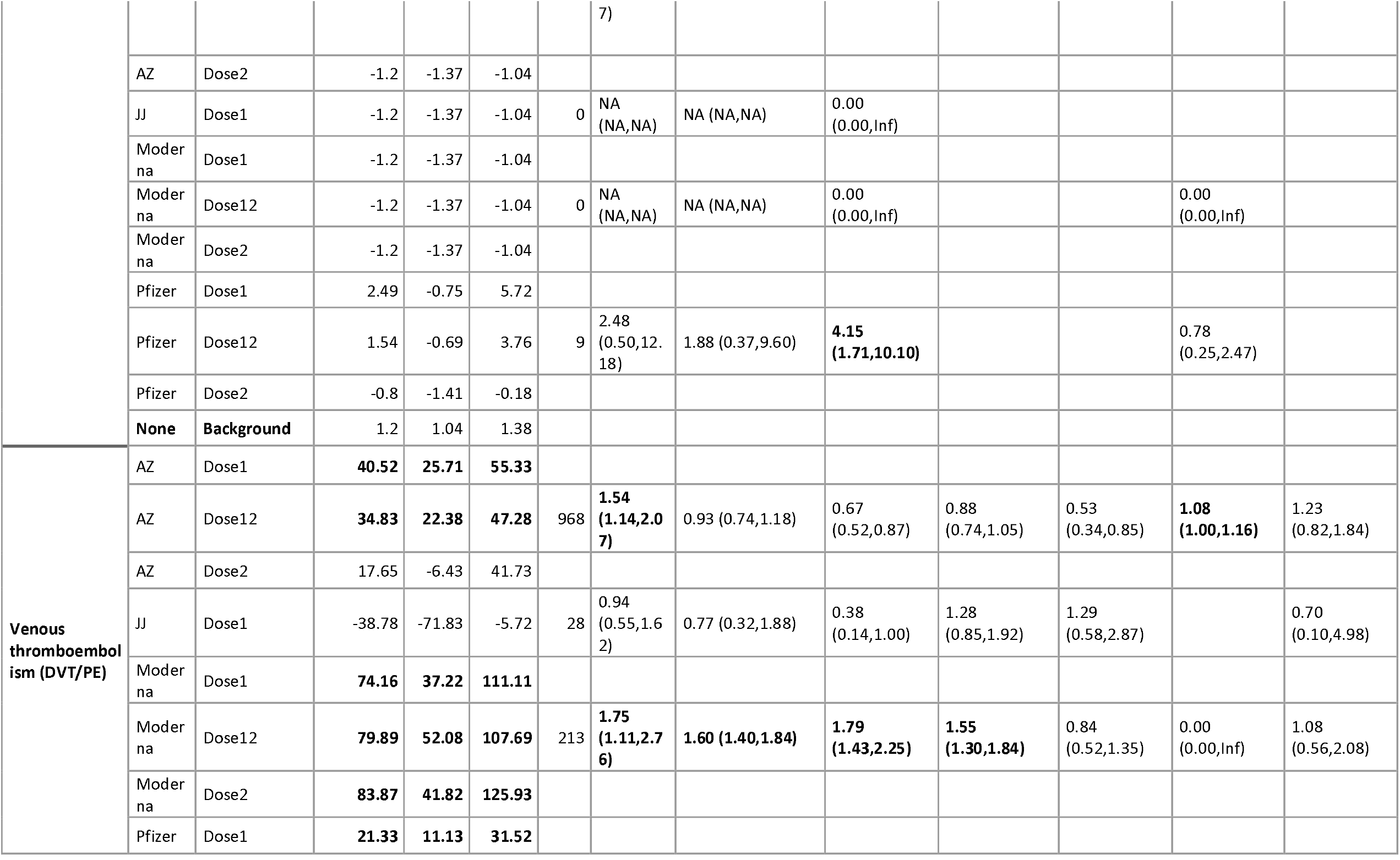
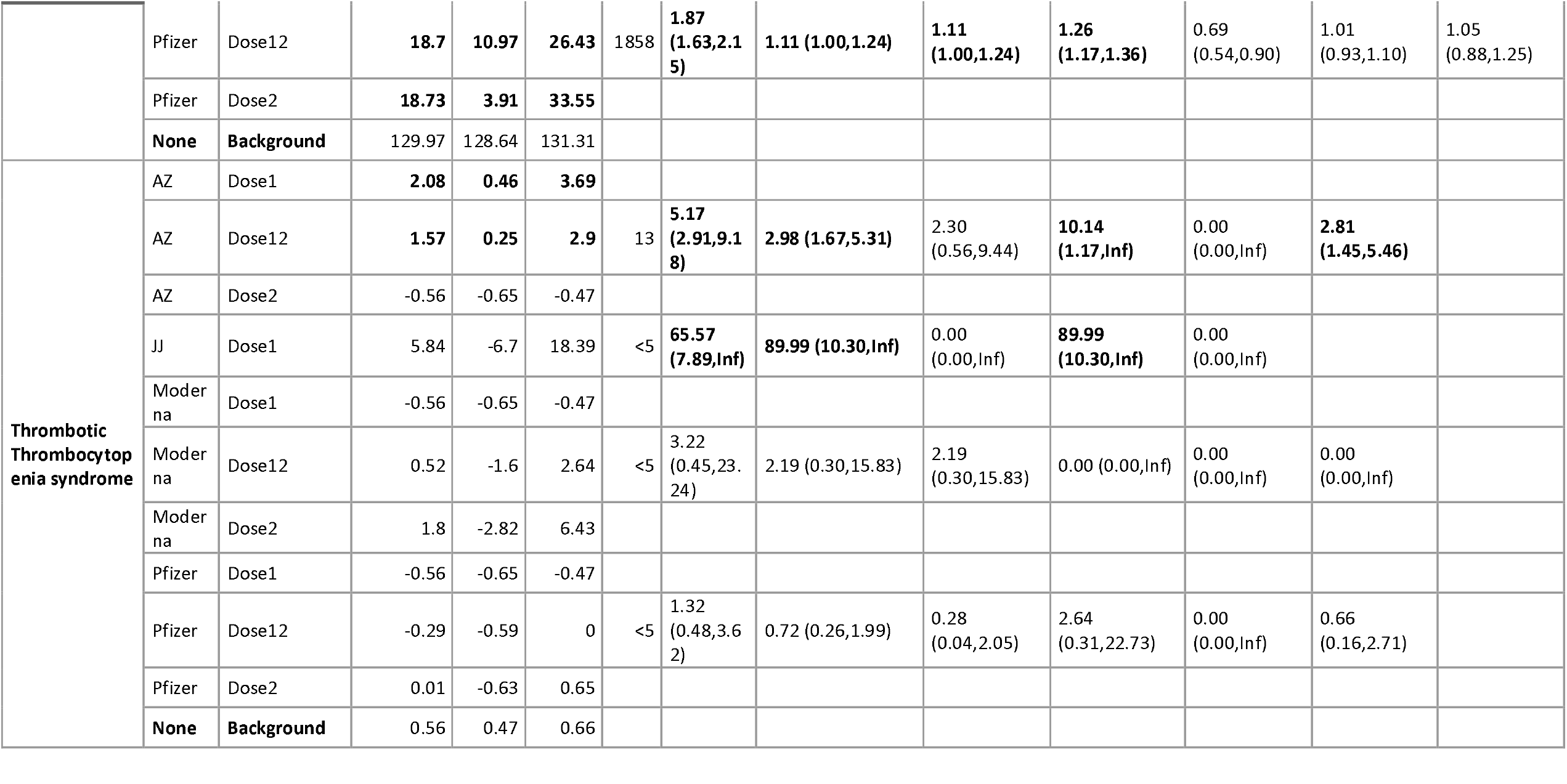
Pooled age-standardized rate differences and incidence rate ratios for AESI following COVID-19 vaccination in the 28 days after dose 1 and/or 2 compared with background rates in 2020.

## Discussion

This study aimed to monitor the safety of the four different covid-19 vaccines that were authorized through the European Medicines Agency and MHRA in 2020-2021. These include two mRNA platform vaccines (Pfizer BioNTech COVID-19 mRNA vaccine BNT162b2 or Comirnaty and Moderna mRNA-1273 COVID-19 vaccine or Spikevax) and two that use an adenovirus vector (ChAdox COVID-19 vaccine / Vaxzevria and COVID-19 vaccine Janssen Ad26. COV2-S [recombinant]). To monitor the safety of these vaccines we used four different health care data sources and more than 25 million people, among them 12.1 million persons had received at least one covid-19 vaccination and were monitored for 29 AESI. The majority of COVID-19 vaccine recipients received either Comirnaty or Vaxzevria, the share of Spikevax (6%) and Janssen (2%) was very low in those data sources at the moment of last data extraction. User patterns differed substantially between the UK and EU countries. Our observations are consistent with the vaccination strategies described by the European Center for Disease Control and Prevention. From the start, vaccinations have been rolled out in phases through various priority groups. Countries initially prioritised elderly people, residents and personnel of long-term care facilities, healthcare workers, social care personnel, and people with certain comorbidities[58]. All EU/EEA countries then opened vaccination to the general population, with all offering vaccination to those aged 12 years and over. In Italy, Vaxzevria and Janssen vaccine are restricted to those 60 years and older, following the signals about (thrombotic)thromboembolic events with the adenovirus platform vaccines, in Spain Comirnaty and Spikevax are recommended for elderly (≥70), pregnant women and individuals with high-risk conditions, and other age groups are according to availability, Vaxzevria should only be used in 60 years and older and Janssen COVID-19 vaccine primarily for those > 40 years of age. In the Netherlands Comirnaty and Spikevax can be used in all people, Vaxzevria is not used anymore and Janssen vaccine only for 18 years and older. Spikevax had a similar user profile as Comirnaty but had very limited use.Janssen vaccine was used by very few people and in Netherlands mostly in young people. The Joint Committee on Vaccination and Immunisation (JCVI) in the UK, for both Pfizer/BioNTech and Oxford/AstraZeneca, the vaccine should first be given to residents in a care home for older adults and their carers then to those over 80 years old as well as frontline health and social care workers, then to the rest of the population in order of age and clinical risk factors. The JCVI also decided that the impact of the second dose is likely to be modest and most of the initial protection from clinical disease is after the first dose of vaccine, they decided that prioritising the first doses of vaccine for as many people as possible on the priority list would protect the greatest number of at-risk people in the shortest possible time this meant that second doses of both vaccines were to be administered towards the end of the recommended vaccine dosing schedule of 12 weeks[59]. Our data reflect the initial strategy with long distances between dose 1 and 2 for both Vaxzevria as well as Comirnaty.

Incidence rates in 2020, were consistent with those observed during the ACCESS project, but lower than 2017-2019 for some cardiac injury events, that are frequent[22]. Incidence rates for the majority of AESI were very rare (<10/100,000 PY) and for those we had limited power to detect elevations of incidence rates post-vaccination. Even when monitoring 12.1 million exposed persons, the risk period is very short: 56 days maximum for 2-dose regimens, and 28 days for a one dose regimen. For adequate and rapid monitoring of such events more data sources should be included.

Several associations were observed in this study most of which have already been part of regulatory discussions. Anaphylaxis was detected immediately after roll out and led to the 15-20 minute waiting time at the immunization sites[5]. Erythema multiforme was investigated by the PRAC for Comirnaty and Spikevax. In the October 2021 meeting PRAC decided that based on the case reports and the fact that there is a plausible mechanism for how the vaccine may cause erythema multiforme, the product information should be updated to include erythema multiforme as a side effect of Spikevax and Comirnaty [60] [61]. In our study we only observed an association of erythema multiforme after Spikevax and not Comirnaty.

In this study we observed an increased rate of GBS following Janssen vaccine based on less than five cases in the post-dose 1 28-day risk interval. Based on case reports and a 42-day risk interval, Woo et al. estimated an observed to expected ratio of 4.18 (95% CI, 3.47-4.98) [62]. The PRAC has reviewed this signal and considered that there might be a causal association[17]. We observed an relevant elevated association between Janssen COVID-19 vaccine and SOCV, cutaneous vasculitis was added as risk to the Janssen vaccine label following several published case reports and review of 37 cases until end of October by PRAC[63]. We found an association between Thrombocytopenia, Janssen vaccine and Spikevax. Thrombocytopenia or low platelets after Vaxzevria and Janssen vaccine have been assessed, and this has been included in the summary of product characteristics[64]. Cases of ITP after Spikevax have been assessed by regulators in July 2021, but a clear causal relationship could not yet be assessed[65]. VTE was associated with Spikevax and Comirnaty in our study, with minor significant elevations, Tobiaqy also described cases based on an analysis of Eudravigilance[52]. We observed and increased excess rate of VTE after Vaxzevria and Janssen vaccine but this disappeared after adjustment for factors associated for age, gender, prior covid-19 and any risk factor for severe covid-19 in our study. We observed relevant increased association between TTS and Janssen COVID-19 and Vaxzevria vaccine in our study consistent with publications and PRAC assessment[9, 66, 67].^2^

Our data are compatible with the findings from the US based Vaccine Safety Datalink which monitored 23 AESI across almost 12 million mRNA COVID-19 vaccine doses (57% Pfizer-BioNTech, 43% Moderna) administered to 6.2 million individuals aged 12 years or older. No outcomes met the prespecified signalling criteria for statistical significance. Rate ratios (RRs) were largest for thrombotic thrombocytopenic purpura (2.60), cerebral venous sinus thrombosis (1.55), and transverse myelitis (1.45), but these measures of association had wide 95% CIs and nonsignificant *P* values. The highest estimates of excess cases per million doses were 7.5 (95% CI, −0.1 to 14.0) for venous thromboembolism, 1.2 (95% CI, −6.9 to 8.3) for acute myocardial infarction, and 1.2 (95% CI, −2.1 to 3.3) for myocarditis/pericarditis[68].

This study is monitoring a large number of vaccines across a wide range of pre-specified AESI, yet this study has several limitations. First of all, the study was designed for monitoring of AESI occurrence and not for causal inference. As part of monitoring, we compared the 28-day post-vaccination period after each dose to the background rate in 2020 rather than a parallel comparator. Since the lockdown has lowered health care seeking behaviour, the 2020 rate may be lower than other years, which we observed for cardiac injury conditions in the ACCESS study[22]. This is why we use an IRR of 2 to consider an association relevant. We adjusted for main risk factors associated with vaccination, but cannot exclude residual confounding as we did not consider individual risk factors for each AESI, neither did we design the study for causal inference. Although comparisons are done within data source, where event recording may stay relatively stable, temporal effects due to awareness (e.g.TTS) and therefore differential misclassification cannot be excluded. There are also limitations due to specifics in the data sources that we use and the data that they capture: ARS data comprises emergency care visits and hospitalizations which explains why the rates of anaphylaxis were higher than in other data sources. It also explains the lower rates of conditions that are not typically seen in this setting such as chilblains and anosmia/ageusia. In BIFAP-PC there could be some misclassification of certain events i.e. more severe cases, since these cases will be better recorded in the hospital setting. However, since BIFAP data in a subset of regions has been linked to hospital diagnoses, we were able to use this PC_HOSP subpopulation to more precisely ascertain AESI cases. Results for both BIFAP subpopulations (PC and PC_HOSP) are generally consistent with those of other DAPS with similar characteristics. Although the study variables have been created in a harmonized manner, there may exist some residual discrepancies in the list of codes among the different coding systems used by the DAPs participating in this study, which may have affected some AESI IRs and lead to some differences with IRs from other data sources. This may be the case of meningoencephalitis, which shows an age pattern slightly different to the rest of DAPs in the oldest age groups in 2020. In addition to this, the lower IRs observed for acute coronary artery disease in BIFAP might also be explained by the fact that lower IRs of ischemic heart disease in Spain compared to those in other European countries has also been seen. The second report on cardiovascular disease (CVD) statistics for the member countries of the European Society of Cardiology (ESC) reported a lower incidence of ischemic heart disease in 2017 in Spain than in Italy, the Netherlands, and the UK[69]. For PHARMO, vaccination status and date of vaccination was obtained from the electronic medical record of the GP and might be incomplete, a large proportion of vaccine brands was missing. The GP Database contains vaccinations administered by GPs and by the public health service, as GPs receive an automated notification when a patient has been vaccinated via the public health service (provided that individuals have given their consent). Vaccines administered at hospitals were missed. In addition, only ICPC coded events were considered in this study. For some events, ICPC codes do not exist. Events that were diagnosed in secondary care were only captured if recorded by the GP. Another limitation is that within the PHARMO data death is under-recorded or delayed. As a result, person time, especially person time in older persons may be overestimated, which leads to an underestimation of the rates. The event rates in this group will be underestimated accordingly.

In CPRD Aurum, only data from primary care were used in this study. Hospital data was not available for the relevant time window. Like in PHARMO, events diagnosed in secondary care were only captured when recorded in the GP practice, which means that some outcomes could have been underdiagnosed. Data from CPRD Aurum ran until the end of May 2021, which means that the number of vaccinations among younger individuals was still relatively low with the vaccination strategy in the UK compared to other DAPs.

## Conclusion

this study showed that we could monitor a large number of AESI across 4 data sources in four countries using the Conception common data model, common analytics and an agile way for semantic harmonization across multiple disease diagnosis vocabularies, data were periodically shared with the EMA through an interactive dashboard. COVID-19 vaccines had different user patterns across the countries in terms of type, distance between dose 1 and 2 and the populations targeted. Several (relevant) elevations of rates were observed, most of which have been part of regulatory discussions such as the haematological events, neurological events and erythema multiforme. This study was not designed for causal inference and hypothesis testing studies would be needed to estimate the excess risk adequately with proper control for risk factors for the outcomes. In spite of the large numbers of vaccinees, power is limited for the events that are rare <10/100,000 PY and continuous monitoring and scaling up is required to monitor this better, or test hypotheses.

## Supporting information

https://www.dropbox.com/s/29lt3d0kyaxehfe/ECVM%20WP2%20Manuscript.docx?dl=0

## Data Availability

The protocol for this study is publicly available
The full report for this study is publicly available at Zenodo. https://doi.org/10.5281/zenodo.6762311

## Acknowledgement

The research leading to these results was conducted as part of the activities of the EU PE&PV (Pharmacoepidemiology and Pharmacovigilance) Research Network (led by Utrecht University) with collaboration from the Vaccine Monitoring Collaboration for Europe network (VAC4EU). Scientific work for this project was coordinated by the University Medical Center Utrecht. The project has received support from the European Medicines Agency under the Framework service contract nr EMA/2018/28/PE. The data pipeline was developed as part of the IMI-ConcePTION project.

This document expresses the opinion of the authors of the paper, and may not be understood or quoted as being made on behalf of or reflecting the position of the European Medicines Agency or one of its committees or working parties.

The authors from BIFAP would like to acknowledge the excellent collaboration of the primary care practitioners and paediatricians, and also the support of the regional authorities participating in the database.

We acknowledge the input from the Vaccine monitoring Collaboration for Europe and its members.

We are grateful to prof. Dr. MJC Eijkemans for statistical support and Susana Perez-Gutthann and Rachel Weinrib for review and comments as well as Dr. Daniel Weibel for VAC4EU secretariat support.

## Objectives

This study aimed to monitor use of COVID-19 vaccines and incidence rates of pre-specified adverse events of special interest (AESI) of COVID-19 vaccines prior to and after COVID-19 vaccination. This study was not aimed to test a specific hypothesis.

## Design

A retrospective cohort study including subjects from January 1, 2020 to October 31^st^, 2021, or latest availability of data.

## Footnotes

1 https://github.com/cran/dsr.

## Notes

### Competing Interest Statement

No author has direct financial conflicts of interest. Authors work for organizations who are conducting research for multiple study requesters including the European Medicines Agency and vaccine manufactures. All studies are conducted according to the ENCePP code of conduct for scientific independence

### Clinical Protocols

https://www.encepp.eu/encepp/viewResource.htm?id=47901

### Author Declarations

This study is based on re-use of clinical data that is pseudonomized. This does not require review from IRBs. Each data access provider (PHARMO, ARS, BIFAP, CPRD) has documented clearance from their respective local governance organizations that oversee re-use of available pseudonomized data for the objectives described.

