## Supplementary material for "Cohort monitoring of 29 Adverse Events of Special Interest prior to and after COVID-19 vaccination in four large European electronic healthcare data sources": https://www.dropbox.com/s/29lt3d0kyaxehfe/ECVM%20WP2%20Manuscript.docx?dl=0

**Supplementary table 1:	Conditions considered to be at-risk conditions for severe COVID-19**

| **At-risk medical conditions identified by diagnosis codes (see ECVM codesheet)** | **Medicinal product proxy(ies) (ATC code)** |
| --- | --- |
| Cancer (with chemo/immuno/radio-therapy, cancer treatment, immunosuppressant; targeted cancer treatment (such as protein kinase inhibitors or PARP inhibitors); blood or bone marrow cancer (such as leukemia, lymphoma, myeloma)) | Alkylating agents (L01A)  Antimetabolites (L01B)  Plant alkaloids and other natural products (L01C)  Cytotoxic antibiotics and related substances (L01D)  Other antineoplastic agents (L01X)  Hormones and related agents (L02A)  Hormone antagonists and related agents (L02B)  Immunostimulants (L03)  Immunosuppressants (L04) |
| Type 1& 2 Diabetes | Blood glucose lowering drugs A10A & A10B |
| Obesity (BMI > 30) | Peripherally acting anti-obesity products (A08AB)  Centrally acting anti-obesity products (A08AA) |
| Cardiovascular disease/ Serious heart conditions including heart failure, coronary artery disease, cardiomyopathies | [A](https://www.whocc.no/atc_ddd_index/?code=C01B&showdescription=yes)ntiarrhythmics, class I and III (C01B)  [Cardiac stimulants excl. Cardiac glycosides](https://www.whocc.no/atc_ddd_index/?code=C01C&showdescription=yes) (C01C)  Vasodilators used in cardiac diseases (C01D)  [Other cardiac preparations](https://www.whocc.no/atc_ddd_index/?code=C01E&showdescription=yes) (C01E)  Antithrombotic agents (B01A)  Antihypertensives (C02, C03, C07, C08, C09) |
| Chronic lung disease including COPD, asthma | Drugs for obstructive airway diseases (R03)  [Lung surfactants](https://www.whocc.no/atc_ddd_index/?code=R07AA&showdescription=yes) (R07AA)  [Respiratory stimulants](https://www.whocc.no/atc_ddd_index/?code=R07AB&showdescription=yes) (R07AB) |
| Chronic kidney disease | Erythropoietin (B03XA01) |
| HIV | Protease inhibitors (J05AE)  Combinations to treat HIV (J05AR)  NRTI (J05AF)  NNRTI (J05AG) |
| Immunosuppression | Immunosuppressants (L04A)  Corticosteroids (H02) |
| Sickle Cell Disease | Hydroxyurea (L01XX05)  Other hematological agents (B06AX) |

Supplementary table 2: Cohort characteristics at study start 1/1/2020, and first dose of any COVID-19 vaccine in IT-ARS

| **Variable** | **Values** | **Baseline 1/1/2020 total population** | | | **Pfizer 1^st^ dose** | | **Moderna 1^st^ dose** | | | AstraZeneca **1^st^ dose** | | **Janssen 1^st^ dose** | |
| --- | --- | --- | --- | --- | --- | --- | --- | --- | --- | --- | --- | --- | --- |
| **Persons 1^st^ dose** | N |  |  | 1,320,326 | | 69.6% | 184,013 | 9.7% | 332,872 | | 17.6% | 58,513 | 3.1% |
| **follow-up 1^st^-2^nd^ dose** | PY |  |  | 87,593 | | 50.5% | 13,726 | 7.9% | 66,133 | | 38.1% | 6,140 | 3.5% |
| **Month 1st vaccination** |  |  |  | 12 | |  | 1 |  | 2 | |  | 3 |  |
| **2021 January** | N |  |  | 62,665 | | 4.7% | 2,984 | 1.6% | 0 | | 0% | 0 | 0% |
| **2021 February** | N |  |  | 68,773 | | 5.2% | 5,938 | 3.2% | 48,174 | | 14.5% | 0 | 0% |
| **2021 March** | N |  |  | 117,022 | | 8.9% | 28,654 | 15.6% | 89,203 | | 26.8% | 1 | 0% |
| **2021 April** | N |  |  | 248,304 | | 18.8% | 26,011 | 14.1% | 110,435 | | 33.2% | 10,250 | 17.5% |
| **2021 May** | N |  |  | 338,400 | | 25.6% | 60,291 | 32.8% | 82,873 | | 24.9% | 24,871 | 42.5% |
| **2021 June** | N |  |  | 483,579 | | 36.6% | 60,135 | 32.7% | 2,187 | | 0.7% | 23,391 | 40% |
| **2020 December** | N |  |  | 1,583 | | 0.1% | 0 | 0% | 0 | | 0% | 0 | 0% |
| **Age in years** | Min | 0 |  | 11 | |  | 9 |  | 1 | |  | 18 |  |
|  | P25 | 29 |  | 48 | |  | 39 |  | 57 | |  | 60 |  |
|  | P50 | 49 |  | 59 | |  | 57 |  | 69 | |  | 62 |  |
|  | Mean | 47 |  | 60 | |  | 52 |  | 63 | |  | 65 |  |
|  | P75 | 66 |  | 76 | |  | 64 |  | 74 | |  | 69 |  |
|  | Max | 119 |  | 108 | |  | 103 |  | 91 | |  | 102 |  |
| **Age in categories** | 0-4 | 113,669 | 3.3% | 0 | | 0% | 0 | 0% | 1 | | 0% | 0 | 0% |
|  | 5-11 | 211,885 | 6.1% | 1 | | 0% | 2 | 0% | 1 | | 0% | 0 | 0% |
|  | 12-17 | 185,91 | 5.3% | 13,924 | | 1.1% | 120 | 0.1% | 0 | | 0% | 0 | 0% |
|  | 18-24 | 212,915 | 6.1% | 61,759 | | 4.7% | 13,613 | 7.4% | 2,004 | | 0.6% | 58 | 0.1% |
|  | 25-29 | 155,684 | 4.5% | 37,391 | | 2.8% | 4,497 | 2.4% | 5,606 | | 1.7% | 78 | 0.1% |
|  | 30-39 | 359,062 | 10.3% | 103,599 | | 7.8% | 30,177 | 16.4% | 15,756 | | 4.7% | 206 | 0.4% |
|  | 40-49 | 521,342 | 14.9% | 148,244 | | 11.2% | 27,111 | 14.7% | 34,948 | | 10.5% | 434 | 0.7% |
|  | 50-59 | 562,496 | 16.1% | 297,275 | | 22.5% | 51,110 | 27.8% | 36,370 | | 10.9% | 7,014 | 12% |
|  | 60-69 | 448,863 | 12.9% | 216,806 | | 16.4% | 33,056 | 18% | 79,766 | | 24% | 37,411 | 63.9% |
|  | 70-79 | 401,694 | 11.5% | 152,143 | | 11.5% | 23,050 | 12.5% | 158,292 | | 47.6% | 13,162 | 22.5% |
|  | 80+ | 316,103 | 9.1% | 289,184 | | 21.9% | 1,277 | 0.7% | 128 | | 0% | 150 | 0.3% |
|  | 60+ | 1,166,660 | 33.4% | 658,133 | | 49.8% | 57,383 | 31.2% | 238,186 | | 71.6% | 50,723 | 86.7% |
| **Person years across sex** | Female | 2,657,824 | 52.1% | 48,204 | | 55% | 6,628 | 48.3% | 37,505 | | 56.7% | 3,244 | 52.8% |
|  | Male | 2,445,817 | 47.9% | 39,389 | | 45% | 7,098 | 51.7% | 28,628 | | 43.3% | 2,896 | 47.2% |
| **At risk population at date of vaccination** | Cardiovascular disease | 969,895 | 27.8% | 620,684 | | 47% | 70,923 | 38.5% | 162,002 | | 48.7% | 28,471 | 48.7% |
|  | Cancer | 84,142 | 2.4% | 57,368 | | 4.3% | 20,967 | 11.4% | 8,821 | | 2.6% | 1,344 | 2.3% |
|  | Chronic lung disease | 195,898 | 5.6% | 123,168 | | 9.3% | 16,360 | 8.9% | 25,816 | | 7.8% | 4,191 | 7.2% |
|  | HIV | 8,728 | 0.3% | 4,228 | | 0.3% | 1,779 | 1% | 389 | | 0.1% | 78 | 0.1% |
|  | Chronic kidney disease | 17,536 | 0.5% | 15,140 | | 1.1% | 4,210 | 2.3% | 877 | | 0.3% | 120 | 0.2% |
|  | Diabetes | 193,969 | 5.6% | 134,732 | | 10.2% | 19,235 | 10.5% | 18,690 | | 5.6% | 3,254 | 5.6% |
|  | Severe obesity | 5,391 | 0.2% | 5,101 | | 0.4% | 1,057 | 0.6% | 589 | | 0.2% | 125 | 0.2% |
|  | Sickle cell disease | 3,56 | 0.1% | 2,504 | | 0.2% | 603 | 0.3% | 185 | | 0.1% | 19 | 0% |
|  | Use of immunosuppressants | 207,855 | 6% | 192,122 | | 14.6% | 33,640 | 18.3% | 42,622 | | 12.8% | 7,761 | 13.3% |
|  | Any risk factors | 1,200,345 | 34.4% | 739,700 | | 56% | 94,645 | 51.4% | 188,759 | | 56.7% | 33,550 | 57.3% |

Supplementary table S3: Cohort characteristics at study start 1/1/2020, and first dose of any COVID-19 vaccine in ES- BIFAP-PC

| Variable | Values | All population at 1/1/2020 |  | Pfizer | | Moderna | | AstraZeneca | | Janssen | | Unknown brand | |
| --- | --- | --- | --- | --- | --- | --- | --- | --- | --- | --- | --- | --- | --- |
| **Persons 1^st^ dose** | N |  |  | 2,808,700 | 70.3% | 447,401 | 11.2% | 537,122 | 13.4% | 201,543 | 5% | 233 | 0% |
| **follow-up 1^st^-2^nd^ dose** | PY |  |  | 176,483 | 47.9% | 36,474 | 9.9% | 115,480 | 31.3% | 40,061 | 10.9% | 30 | 0% |
| Month of first vaccination |  |  |  | 12 |  | 1 |  | 1 |  | 2 |  | 12 |  |
| 2021 January | N |  |  | 165,885 | 5.9% | 4,263 | 1% | 2 | 0% | 0 | 0% | 33 | 14.2% |
| 2021 February | N |  |  | 103,859 | 3.7% | 18,075 | 4% | 23,169 | 4.3% | 2 | 0% | 29 | 12.4% |
| 2021 March | N |  |  | 191,809 | 6.8% | 26,920 | 6% | 100,409 | 18.7% | 4 | 0% | 40 | 17.2% |
| 2021 April | N |  |  | 520,783 | 18.5% | 50,584 | 11.3% | 230,122 | 42.8% | 14,564 | 7.2% | 29 | 12.4% |
| 2021 May | N |  |  | 351,643 | 12.5% | 129,507 | 28.9% | 150,394 | 28% | 37,012 | 18.4% | 47 | 20.2% |
| 2021 June | N |  |  | 763,483 | 27.2% | 51,301 | 11.5% | 18,876 | 3.5% | 110,723 | 54.9% | 25 | 10.7% |
| 2021 July | N |  |  | 382,209 | 13.6% | 103,590 | 23.2% | 13,075 | 2.4% | 34,312 | 17% | 21 | 9% |
| 2021 August | N |  |  | 316,591 | 11.3% | 63,161 | 14.1% | 1,075 | 0.2% | 4,926 | 2.4% | 5 | 2.1% |
| 2020 December | N |  |  | 12,438 | 0.4% | 0 | 0% | 0 | 0% | 0 | 0% | 4 | 1.7% |
| Age in years | Min | 0 |  | 2 |  | 3 |  | 5 |  | 9 |  | 13 |  |
|  | P25 | 27 |  | 39 |  | 30 |  | 59 |  | 42 |  | 30 |  |
|  | P50 | 46 |  | 51 |  | 49 |  | 61 |  | 49 |  | 47 |  |
|  | Mean | 45 |  | 53 |  | 47 |  | 58 |  | 50 |  | 50 |  |
|  | P75 | 63 |  | 71 |  | 58 |  | 64 |  | 56 |  | 64 |  |
|  | Max | 113 |  | 112 |  | 103 |  | 101 |  | 102 |  | 102 |  |
| Age in categories | 0-4 | 220,67 | 3.8% | 10 | 0% | 2 | 0% | 0 | 0% | 0 | 0% | 0 | 0% |
|  | 5-11 | 385,632 | 6.6% | 1,550 | 0.1% | 278 | 0.1% | 3 | 0% | 1 | 0% | 0 | 0% |
|  | 12-17 | 335,254 | 5.8% | 168,143 | 6% | 25,117 | 5.6% | 102 | 0% | 59 | 0% | 4 | 1.7% |
|  | 18-24 | 365,851 | 6.3% | 142,276 | 5.1% | 53,718 | 12% | 11,704 | 2.2% | 5,049 | 2.5% | 37 | 15.9% |
|  | 25-29 | 278,046 | 4.8% | 109,504 | 3.9% | 27,754 | 6.2% | 11,452 | 2.1% | 3,250 | 1.6% | 15 | 6.4% |
|  | 30-39 | 711,513 | 12.2% | 315,236 | 11.2% | 65,785 | 14.7% | 28,665 | 5.3% | 12,150 | 6% | 33 | 14.2% |
|  | 40-49 | 939,792 | 16.2% | 566,889 | 20.2% | 53,176 | 11.9% | 39,664 | 7.4% | 85,463 | 42.4% | 32 | 13.7% |
|  | 50-59 | 880,812 | 15.1% | 526,549 | 18.7% | 119,088 | 26.6% | 63,424 | 11.8% | 59,002 | 29.3% | 41 | 17.6% |
|  | 60-69 | 702,145 | 12.1% | 197,008 | 7% | 36,512 | 8.2% | 381,833 | 71.1% | 25,465 | 12.6% | 22 | 9.4% |
|  | 70-79 | 531,479 | 9.1% | 435,772 | 15.5% | 36,625 | 8.2% | 241 | 0% | 10,614 | 5.3% | 11 | 4.7% |
|  | 80+ | 465,361 | 8% | 345,763 | 12.3% | 29,346 | 6.6% | 34 | 0% | 490 | 0.2% | 38 | 16.3% |
|  | 60+ | 1,698,985 | 29.2% | 978,543 | 34.8% | 102,483 | 23% | 382,108 | 71.1% | 36,569 | 18.1% | 71 | 30.4% |
| Person years across sex | Female | 4,767,360 | 51.1 | 92,692 | 52.5% | 19,026 | 52.2% | 63,238 | 54.8% | 18,408 | 45.9% | 19 | 63.3% |
|  | Male | 4,569,396 | 48.9 | 83,791 | 47.5% | 17,448 | 47.8% | 52,241 | 45.2% | 21,653 | 54.1% | 11 | 36.7% |
| At risk population at date of vaccination | Cardiovascular disease | 1,107,931 | 19% | 851,851 | 30.3% | 105,818 | 23.7% | 174,031 | 32.4% | 38,866 | 19.3% | 64 | 27.5% |
|  | Cancer | 67,793 | 1.2% | 66,405 | 2.4% | 13,867 | 3.1% | 11,595 | 2.2% | 3,158 | 1.6% | 1 | 0.4% |
|  | Chronic lung disease | 248,979 | 4.3% | 224,775 | 8% | 30,356 | 6.8% | 38,438 | 7.2% | 12,181 | 6% | 21 | 9% |
|  | HIV | 702 | 0% | 749 | 0% | 238 | 0.1% | 131 | 0% | 64 | 0% | 0 | 0% |
|  | Chronic kidney disease | 16,539 | 0.3% | 22,730 | 0.8% | 2,447 | 0.5% | 2,169 | 0.4% | 536 | 0.3% | 0 | 0% |
|  | Diabetes | 323,509 | 5.6% | 252,396 | 9% | 31,789 | 7.1% | 56,447 | 10.5% | 12,571 | 6.2% | 12 | 5.2% |
|  | Severe obesity | 47,823 | 0.8% | 49,821 | 1.8% | 6,567 | 1.5% | 10,447 | 1.9% | 3,835 | 1.9% | 2 | 0.9% |
|  | Sickle cell disease | 2,882 | 0% | 2,373 | 0.1% | 413 | 0.1% | 312 | 0.1% | 113 | 0.1% | 0 | 0% |
|  | Use of immunosuppressants | 70,646 | 1.2% | 84,441 | 3% | 13,704 | 3.1% | 14,889 | 2.8% | 4,831 | 2.4% | 4 | 1.7% |
|  | Any risk factors | 1,392,185 | 23.9% | 1,076,081 | 38.3% | 143,137 | 32% | 223,772 | 41.7% | 56,637 | 28.1% | 80 | 34.3% |

Supplementary table S4 Cohort characteristics at study start 1/1/2020, and first dose of any COVID-19 vaccine in NL-PHARMO

| Variable | Values | total population at 1/1/2020 | | Pfizer | | Moderna | | AstraZeneca | | Janssen | | Unknown manufacturer | |
| --- | --- | --- | --- | --- | --- | --- | --- | --- | --- | --- | --- | --- | --- |
| Persons with a first dose | N |  |  | 568,119 | 67.6% | 67,689 | 8.1% | 68,655 | 8.2% | 22,455 | 2.7% | 113,201 | 13.5% |
| Person-years of follow-up 1^st^ and 2^nd^ dose | PY |  |  | 59,305 | 59.3% | 5,551 | 5.6% | 14,211 | 14.2% | 1,603 | 1.6% | 19,309 | 19.3% |
| Month of first vaccination |  |  |  | 1 |  | 1 |  | 1 |  | 1 |  | 1 |  |
| 2021 January | N |  |  | 465 | 0.1% | 415 | 0.6% | 8 | 0% | 1 | 0% | 1,113 | 1% |
| 2021 February | N |  |  | 1,570 | 0.3% | 520 | 0.8% | 5,769 | 8.4% | 1 | 0% | 7,132 | 6.3% |
| 2021 March | N |  |  | 29,649 | 5.2% | 991 | 1.5% | 13,291 | 19.4% | 7 | 0% | 14,735 | 13% |
| 2021 April | N |  |  | 106,777 | 18.8% | 876 | 1.3% | 21,970 | 32% | 123 | 0.5% | 34,916 | 30.8% |
| 2021 May | N |  |  | 139,336 | 24.5% | 21,236 | 31.4% | 19,590 | 28.5% | 339 | 1.5% | 30,832 | 27.2% |
| 2021 June | N |  |  | 185,136 | 32.6% | 20,465 | 30.2% | 7,006 | 10.2% | 12,863 | 57.3% | 18,977 | 16.8% |
| 2021 July | N |  |  | 104,323 | 18.4% | 23,152 | 34.2% | 1,021 | 1.5% | 9,116 | 40.6% | 5,493 | 4.9% |
| 2021 August | N |  |  | 863 | 0.2% | 34 | 0.1% | 0 | 0% | 5 | 0% | 3 | 0% |
| Age in years | Min | 0 |  | 2 |  | 15 |  | 7 |  | 17 |  | 1 |  |
|  | P25 | 23 |  | 37 |  | 36 |  | 60 |  | 21 |  | 53 |  |
|  | P50 | 44 |  | 56 |  | 47 |  | 62 |  | 26 |  | 62 |  |
|  | Mean | 43 |  | 54 |  | 45 |  | 61 |  | 32 |  | 59 |  |
|  | P75 | 61 |  | 71 |  | 54 |  | 63 |  | 45 |  | 66 |  |
|  | Max | 120 |  | 105 |  | 108 |  | 103 |  | 95 |  | 105 |  |
| Age in categories | 0-4 | 98,505 | 4.3% | 1 | 0% | 0 | 0% | 0 | 0% | 0 | 0% | 8 | 0% |
|  | 5-11 | 169,465 | 7.3% | 16 | 0% | 0 | 0% | 2 | 0% | 0 | 0% | 18 | 0% |
|  | 12-17 | 159,05 | 6.9% | 25,858 | 4.6% | 140 | 0.2% | 18 | 0% | 298 | 1.3% | 866 | 0.8% |
|  | 18-24 | 193,115 | 8.4% | 35,607 | 6.3% | 7,299 | 10.8% | 500 | 0.7% | 10,136 | 45.1% | 4,927 | 4.4% |
|  | 25-29 | 136,145 | 5.9% | 26,007 | 4.6% | 3,769 | 5.6% | 370 | 0.5% | 2,870 | 12.8% | 2,863 | 2.5% |
|  | 30-39 | 270,731 | 11.7% | 66,433 | 11.7% | 9,351 | 13.8% | 1,011 | 1.5% | 2,665 | 11.9% | 6,036 | 5.3% |
|  | 40-49 | 295,905 | 12.8% | 71,856 | 12.6% | 19,666 | 29.1% | 1,821 | 2.7% | 1,456 | 6.5% | 8,457 | 7.5% |
|  | 50-59 | 349,615 | 15.1% | 109,862 | 19.3% | 24,485 | 36.2% | 3,617 | 5.3% | 4,697 | 20.9% | 16,101 | 14.2% |
|  | 60-69 | 302,116 | 13.1% | 73,675 | 13% | 1,530 | 2.3% | 59,144 | 86.1% | 230 | 1% | 52,002 | 45.9% |
|  | 70-79 | 226,903 | 9.8% | 122,296 | 21.5% | 662 | 1% | 841 | 1.2% | 70 | 0.3% | 14,239 | 12.6% |
|  | 80+ | 110,904 | 4.8% | 36,508 | 6.4% | 787 | 1.2% | 1,331 | 1.9% | 33 | 0.1% | 7,684 | 6.8% |
|  | 60+ | 639,923 | 27.7% | 232,479 | 40.9% | 2,979 | 4.5% | 61,316 | 89.2% | 333 | 1.4% | 73,925 | 65.3% |
| Person years across sex | Female | 1,580,705 | 50.7 | 30,548 | 51.5% | 2,667 | 48% | 7,032 | 49.5% | 627 | 39.1% | 10,863 | 56.3% |
|  | Male | 1,536,850 | 49.3 | 28,758 | 48.5% | 2,884 | 52% | 7,179 | 50.5% | 976 | 60.9% | 8,446 | 43.7% |
| At risk population at date of vaccination | Cardiovascular disease | 452,131 | 19.6% | 190,844 | 33.6% | 11,025 | 16.3% | 28,541 | 41.6% | 1,177 | 5.2% | 39,949 | 35.3% |
|  | Cancer | 51,417 | 2.2% | 29,099 | 5.1% | 2,527 | 3.7% | 4,040 | 5.9% | 165 | 0.7% | 6,168 | 5.4% |
|  | Chronic lung disease | 144,273 | 6.2% | 60,974 | 10.7% | 6,081 | 9% | 8,882 | 12.9% | 547 | 2.4% | 12,633 | 11.2% |
|  | HIV | 2,519 | 0.1% | 1,076 | 0.2% | 193 | 0.3% | 189 | 0.3% | 16 | 0.1% | 185 | 0.2% |
|  | Chronic kidney disease | 13,093 | 0.6% | 9,155 | 1.6% | 278 | 0.4% | 1,041 | 1.5% | 16 | 0.1% | 2,494 | 2.2% |
|  | Diabetes | 110,086 | 4.8% | 48,399 | 8.5% | 3,438 | 5.1% | 7,864 | 11.5% | 237 | 1.1% | 9,977 | 8.8% |
|  | Severe obesity | 3,703 | 0.2% | 3,015 | 0.5% | 385 | 0.6% | 1,025 | 1.5% | 57 | 0.3% | 672 | 0.6% |
|  | Sickle cell disease | 827 | 0% | 242 | 0% | 46 | 0.1% | 28 | 0% | 2 | 0% | 52 | 0% |
|  | Use of immunosuppressants | 68,202 | 2.9% | 37,848 | 6.7% | 3,403 | 5% | 5,468 | 8% | 238 | 1.1% | 9,039 | 8% |
|  | Any risk factors | 605,829 | 26.2% | 251,069 | 44.2% | 19,526 | 28.8% | 37,119 | 54.1% | 2,103 | 9.4% | 54,226 | 47.9% |

Supplementary table S5 Cohort characteristics at study start 1/1/2020, and first dose of any COVID-19 vaccine in UK-CPRD

| Variable | Values | Total population at 1/1/2020 | | Pfizer | | Moderna | | AstraZeneca |  |
| --- | --- | --- | --- | --- | --- | --- | --- | --- | --- |
| Persons with a first dose | N |  |  | 1,801,355 | 32.8% | 27,023 | 0.5% | 3,671,672 | 66.8% |
| Person-years of follow-up between first and second dose | PY |  |  | 351,696 | 37.2% | 1,172 | 0.1% | 592,811 | 62.7% |
| Month of first vaccination |  |  |  | 12 |  | 1 |  | 12 |  |
| 2021 January | N |  |  | 855,054 | 47.5% | 10 | 0% | 508,269 | 13.8% |
| 2021 February | N |  |  | 649,482 | 36.1% | 9 | 0% | 1,077,156 | 29.3% |
| 2021 March | N |  |  | 71,747 | 4% | 9 | 0% | 1,669,343 | 45.5% |
| 2021 April | N |  |  | 32,454 | 1.8% | 22,981 | 85% | 326,638 | 8.9% |
| 2021 May | N |  |  | 7,684 | 0.4% | 4,014 | 14.9% | 90,114 | 2.5% |
| 2020 December | N |  |  | 184,934 | 10.3% | 0 | 0% | 152 | 0% |
| Age in years | Min | 0 |  | 1 |  | 16 |  | 4 |  |
|  | P25 | 23 |  | 50 |  | 44 |  | 47 |  |
|  | P50 | 40 |  | 65 |  | 46 |  | 56 |  |
|  | Mean | 41 |  | 62 |  | 45 |  | 56 |  |
|  | P75 | 58 |  | 77 |  | 48 |  | 66 |  |
|  | Max | 120 |  | 111 |  | 92 |  | 110 |  |
| Age in categories | 0-4 | 698,613 | 5% | 10 | 0% | 0 | 0% | 1 | 0% |
|  | 5-11 | 1,137,333 | 8.1% | 17 | 0% | 0 | 0% | 13 | 0% |
|  | 12-17 | 914,983 | 6.5% | 12,412 | 0.7% | 25 | 0.1% | 3,843 | 0.1% |
|  | 18-24 | 1,124,457 | 8% | 47,584 | 2.6% | 551 | 2% | 88,712 | 2.4% |
|  | 25-29 | 958,862 | 6.8% | 50,177 | 2.8% | 532 | 2% | 84,502 | 2.3% |
|  | 30-39 | 2,075,853 | 14.7% | 134,641 | 7.5% | 531 | 2% | 277,494 | 7.6% |
|  | 40-49 | 1,940,625 | 13.8% | 199,681 | 11.1% | 24,619 | 91.1% | 713,969 | 19.4% |
|  | 50-59 | 1,965,633 | 13.9% | 294,015 | 16.3% | 564 | 2.1% | 1,064,781 | 29% |
|  | 60-69 | 1,446,722 | 10.3% | 322,598 | 17.9% | 155 | 0.6% | 771,523 | 21% |
|  | 70-79 | 1,129,563 | 8% | 378,051 | 21% | 35 | 0.1% | 522,022 | 14.2% |
|  | 80+ | 708,882 | 5% | 362,169 | 20.1% | 11 | 0% | 144,812 | 3.9% |
|  | 60+ | 3,285,167 | 23.3% | 1,062,818 | 59% | 201 | 0.7% | 1,438,357 | 39.1% |
| Person years across sex | Female | 8,887,890 | 49.9 | 204,369 | 58.1% | 526 | 44.9% | 312,481 | 52.7% |
|  | Male | 8,937,724 | 50.1 | 147,327 | 41.9% | 646 | 55.1% | 280,330 | 47.3% |
| At risk population at date of vaccination | Cardiovascular disease | 2,263,129 | 16% | 851,748 | 47.3% | 1,856 | 6.9% | 1,137,992 | 31% |
|  | Cancer | 165,691 | 1.2% | 94,608 | 5.3% | 139 | 0.5% | 110,744 | 3% |
|  | Chronic lung disease | 931,935 | 6.6% | 281,204 | 15.6% | 1,494 | 5.5% | 452,459 | 12.3% |
|  | HIV | 3,923 | 0% | 1,504 | 0.1% | 5 | 0% | 2,614 | 0.1% |
|  | Chronic kidney disease | 23,076 | 0.2% | 15,143 | 0.8% | 1 | 0% | 13,872 | 0.4% |
|  | Diabetes | 650,872 | 4.6% | 255,170 | 14.2% | 138 | 0.5% | 327,116 | 8.9% |
|  | Severe obesity | 87,926 | 0.6% | 34,650 | 1.9% | 239 | 0.9% | 60,182 | 1.6% |
|  | Sickle cell disease | 2,23 | 0% | 1,065 | 0.1% | 0 | 0% | 1,066 | 0% |
|  | Use of immunosuppressants | 53,977 | 0.4% | 26,324 | 1.5% | 46 | 0.2% | 37,658 | 1% |
|  | Any risk factors | 3,111,051 | 22.1% | 1,068,639 | 59.3% | 3,600 | 13.3% | 1,547,474 | 42.1% |
